# Mortality risk and burden associated with hydroclimate whiplash in the United States

**DOI:** 10.64898/2026.08.18.26360736

**Authors:** Pin Wang, Yiqun Ma, Jennifer D. Stowell, Azar M. Abadi

## Abstract

Hydroclimate whiplash, defined as the rapid transition between unusually wet and dry conditions, is expected to intensify under climate change, yet its population health impacts remain largely unknown. Here we quantified the association between hydroclimate whiplash and mortality across the contiguous United States from 2003 to 2023 using monthly county-level mortality records, standardized precipitation evapotranspiration index data, and two-stage time-series models. We identified overall and direction-specific dry-to-wet and wet-to-dry whiplash events at seasonal and sub-annual timescales and across 5-, 10-, and 20-year recurrence intervals. More severe whiplash events were associated with higher all-cause mortality risk; 5-, 10-, and 20-year sub-annual overall whiplash events increased mortality risk over five months by 3.4%, 4.5%, and 5.7%, respectively. Elevated risks were observed across cause-specific mortality outcomes, with the strongest association for infectious diseases. We estimated that 103,471 deaths were attributable to overall whiplash during the study period. These findings identify hydroclimate whiplash as an emerging climate-related public health threat and suggest that adaptation strategies focused on single hazards may underestimate the health burden of rapid, sequential hydroclimatic extremes.

## Introduction

Anthropogenic climate change is associated not only with rising global mean temperatures but also with increasing climate variability,^1,2^ increasing the frequency and intensity of isolated extreme weather events, including heavy precipitation, floods, and drought, as well as compound events,^3^ commonly defined as climate hazards that spatially and/or temporally interact to produce amplified societal and environmental impacts. Compared with the increasing occurrence of temperature extremes driven by a warming mean climate, changes in precipitation patterns are more strongly governed by climate variability.^4^ Currently, no consensus has been reached on the hydrological trend in response to global warming: one hypothesis projects that wet regions will become wetter while dry regions will become drier, whereas another posits a more generalized long-term aridification of terrestrial areas.^5^

Nevertheless, there is mounting evidence documenting an increase in the rapid alternation between wet and dry conditions, commonly referred to as hydroclimate whiplash, in a changing climate,^6,7^ despite differences in terminology and methods across studies.^8^ A recent study observed a considerable increase in the frequency of historical whiplash events since 1950 and projected a further 52%–113% increase in frequency under a 3°C warming scenario.^7^ Another study projected a 25%–60% increase in frequency and a 30%–100% increase in intensity in semi-arid regions of the globe by the end of this century.^9^ Furthermore, anthropogenic greenhouse gas emissions are projected to increase the risk of hydroclimate whiplash by about 55% by 2079.^10^

These whiplash events are essentially sequential compound hazards that pose a plausible risk to human health. Both floods and drought are extreme weather events with devastating impacts on human society through multiple pathways, including reduced crop yields, deteriorating water availability and quality, declining health, and displacement and forced migration.^11^ Furthermore, compound extreme events can trigger cascading or synergistic health consequences whose magnitude exceeds that of the impacts expected from individual hazards occurring in isolation.^12–14^ This compound effect is supported by our previous study, which found that floods following prior drought, particularly long-term drought, were consistently more strongly associated with childhood diarrhea.^15^ Moreover, drought-to-flood whiplash has been found to increase the abundance of the West Nile virus vector in rural California, suggesting an elevated risk of mosquito-borne diseases.^16^ This hydroclimatic transition is also likely linked to increased incidence of coccidioidomycosis.^17^

In addition, studies have shown that hydroclimate whiplash can amplify wildfire risk at regional to global scales by generating large intra- and interannual variations in biomass through rapid transitions from unusually wet growing seasons to unusually dry fire seasons.^18^ As climate change is expected to intensify hydroclimate variability that could further elevate wildfire risk and associated exposures, the future health burden associated with wildfire smoke may be even greater than current projections suggest.^19^

These interacting extremes may introduce cascading health risks via critical infrastructure failures and service disruptions.^20^ Whiplash events representing rapid shifts between these extremes leave little time for communities and infrastructure to recover, rebuild, and adjust between shocks, thereby potentially undermining institutional and social adaptive capacity and contributing to disproportionate adverse health outcomes.^21^ Currently, hydroclimate whiplash events are still largely conceptualized and described as cascading risks in the literature; to the best of our knowledge, there is no epidemiological evidence on whether, or how, they are associated with human health. Therefore, this study aimed to: (1) examine the association between hydroclimate whiplash and mortality in the contiguous United States; (2) explore whether the association varied across subpopulations and causes of death; and (3) estimate the number and fraction of deaths attributable to whiplash events across space and time.

## Methods

### Data

We obtained mortality data for 2003–2023 from the National Center for Health Statistics (NCHS) at the U.S. Centers for Disease Control and Prevention (U.S. CDC) and aggregated the all-cause and cause-specific death counts at the county-month level. We obtained the monthly standardized precipitation evapotranspiration index (SPEI) at a spatial resolution of 5 km × 5 km for 1895– 2023 from the National Oceanic and Atmospheric Administration’s (NOAA) National Centers for Environmental Information. The SPEI measures drought severity using the standardized climatic water balance between precipitation and potential evapotranspiration, making it sensitive to both precipitation variability and temperature-driven evaporative demand.^22^ We used 3- and 6-month SPEI timescales (hereafter SPEI-3 and SPEI-6) to represent rapid seasonal oscillations and longer-term sub-annual shifts associated with hydroclimate whiplash events, respectively. These timescales are further explained in Text S1.

The urban-rural classification scheme for 2013 was obtained from the NCHS. Monthly mean maximum and minimum temperatures and total precipitation at a resolution of 1 km × 1 km were obtained from the Daily Surface Weather Data on a 1-km Grid for North America, version 4 (Daymet). Monthly mean PM_2.5_ concentration at the same resolution was obtained from the Atmospheric Composition Analysis Group at Washington University in St. Louis. County-level median household income and social vulnerability index (SVI) were obtained from the U.S. Department of Agriculture Economic Research Service and the CDC’s Agency for Toxic Substances and Disease Registry, respectively. SVI incorporates demographic and socioeconomic factors that negatively impact a community’s health resilience.^23^ The total and age-specific county-level population in 2020 (the most recent census) was obtained from the U.S. Census Bureau. We also downloaded the 1 km Köppen-Geiger data from Beck et al.^24^ and assigned to each county with the most common climate across all grid cells within.

### Hydroclimate whiplash identification

We calculated the difference in SPEI between each month and the immediately preceding month during 2003–2023, with positive values indicating a shift toward wetter conditions and negative values indicating a shift toward drier conditions. We identified a hydroclimate whiplash event when the consecutive-month difference crossed a prespecified threshold.^7^ Thresholds were determined according to baseline period, whiplash event type, and recurrence interval (RI). We used the period 1895–1949 as the baseline, representing a relatively stable climate period prior to rapid anthropogenic warming and the substantial observed increase in whiplash frequency starting in the mid-twentieth century.^7^

We considered three whiplash types: overall, dry-to-wet, and wet-to-dry. Overall whiplash was identified as an unusually large absolute change in SPEI, regardless of direction. Directional whiplash was further classified based on the sign of the SPEI change: strongly positive (dry-to-wet) or strongly negative (wet-to-dry), using upper or lower thresholds of the distribution of the differences (see the formula below). These event types capture two distinct transition patterns: interstate transitions across hydroclimate states (e.g., wet to dry) and intrastate changes within the same state, including intensification and recovery. Whiplash identification, event types, and transition patterns are further illustrated in Figure 1.

**Figure 1.**
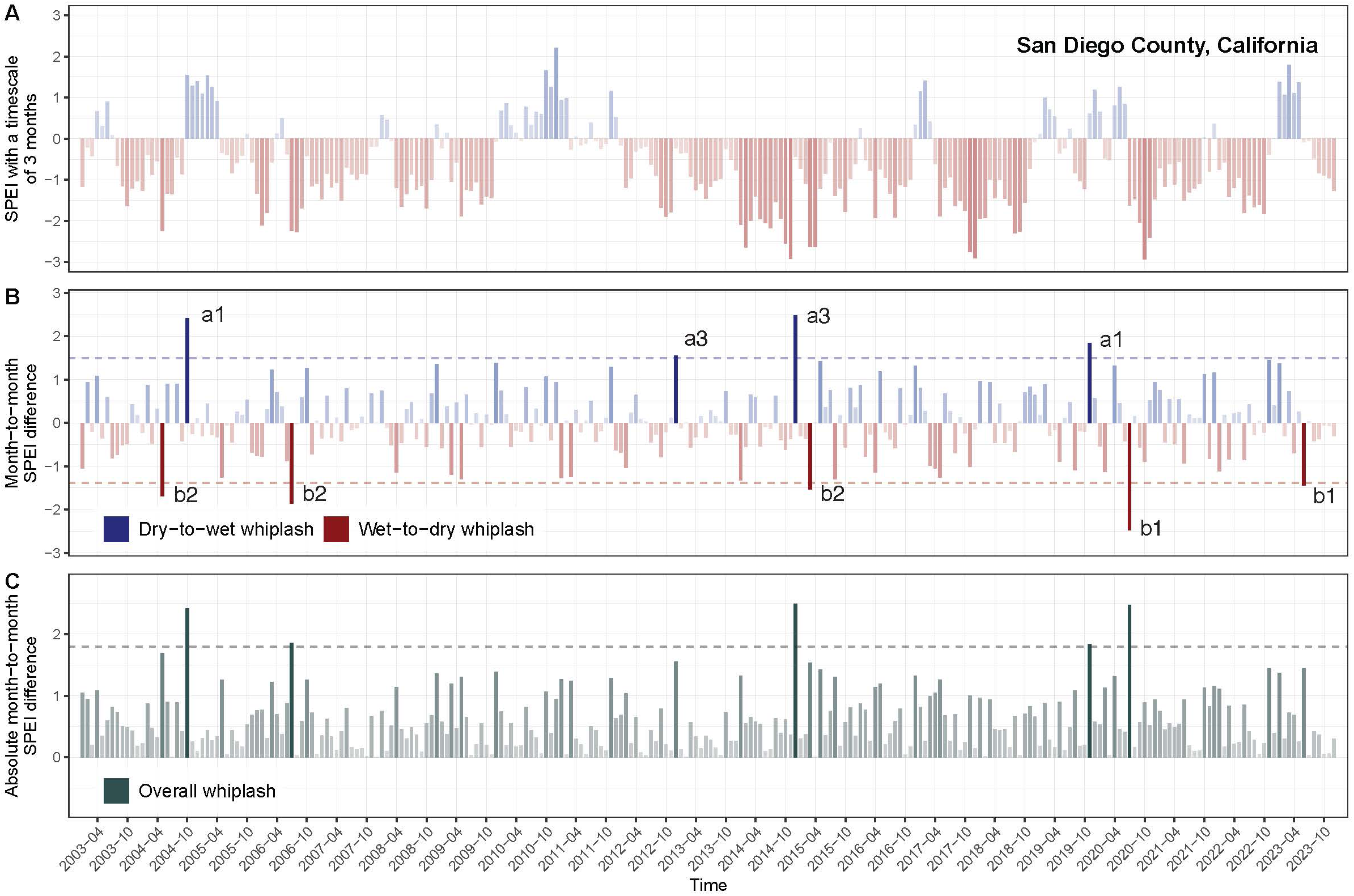
Illustration of hydroclimate whiplash with a 5-year recurrence interval in San Diego County, California during 2003–2023 (A: time-series of SPEI with a timescale of three months; B: time-series of month-to-month SPEI differences; C: time-series of absolute month-to-month SPEI differences. Dark blue and dark red bars represent direction-specific whiplash events, which can be further classified into six patterns: a1. Interstate dry-to-wet transition; a2. Intrastate intensification within the wet state; a3. Intrastate recovery within the dry state; b1. Interstate wet- to-dry transition; b2. Intrastate intensification within the dry state; and b3. Intrastate recovery within the wet state. Types a2 and b3 are not present in this illustration. Dark green bars represent overall whiplash events. Dashed lines denote the thresholds to identify a relevant type of whiplash. SPEI: standardized precipitation evapotranspiration index).

We adopted RIs of 5, 10, and 20 years to represent increasing event severity. The RI, also known as the return period, denotes the average interval between events exceeding a given magnitude; longer RIs correspond to rarer and typically more severe events. Importantly, RI reflects probability rather than periodicity; for example, a 5-year RI corresponds to a 20% chance of occurrence in any given year (or ∼1.67% in any given month), rather than implying that such an event occurs regularly every 5 years.

For month *t* in grid cell *i*, we calculated the consecutive-month SPEI difference and identified a whiplash event *W_i,t_* as:

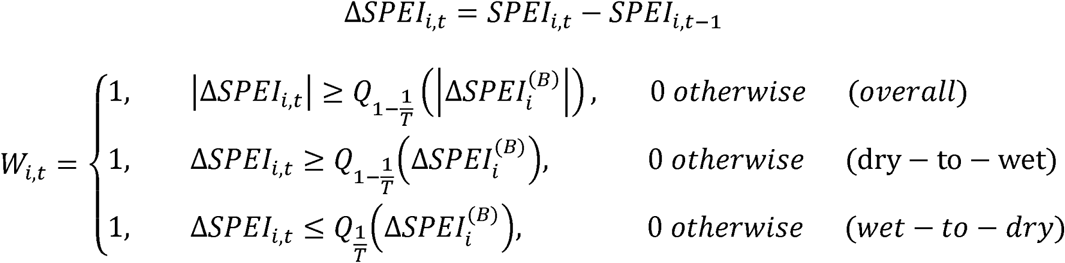

Where *Q_p_*denotes the *p*-th quantile calculated from the baseline period *B*, and *T* represents the RI converted to months, such that *T* = 60 for a 5-year RI. To ensure stable estimates across a gradient of event severity, we applied a set of increasingly stringent thresholds corresponding to different RIs. These thresholds are nested, meaning that more severe events are a subset of less severe ones, rather than forming separate categories.

We then aggregated gridded whiplash data to the county level using a population-weighted approach, in which grid cells were weighted by their resident population within each county. For each county-month, we calculated the fraction of the county population residing in grid cells experiencing whiplash. Counties were classified as exposed when at least 25% of the population was exposed in a given month. This threshold was selected to balance epidemiological relevance and statistical power.

### Statistical analysis

We applied a two-stage time-series model framework to estimate the association between whiplash and mortality risk, as well as the attributable burden. First, to account for conventional exposure-specific and additional lag-specific associations, a standard quasi-Poisson regression with a distributed lag non-linear model was employed in each county to derive county-specific whiplash-mortality associations, adjusting for splines of mean temperature, precipitation, and month with three degrees of freedom (*df*), PM_2.5_, and a factor of year. Specifically, a linear function was used for the exposure-response association, and a natural spline function with four *df* was used for the lag-response association. We predefined a maximum lag period of five months to explore the extended relationship. Then, we pooled the county-specific estimates using a multivariate meta-regression with a combination of county-specific meta-predictors: the proportion of people aged 65 or over, median household income, urban-rural classification, and the SVI.

We used the maximum observed lag to extract the cumulative association, reported as the relative risk (RR). We further estimated the county-month-specific fraction (AF) and number (AN) of deaths attributable to whiplash events using:

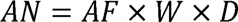

Where

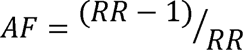

and W and D represent the binary whiplash indicator and total number of deaths in a given county and month, respectively. To capture the mortality burden across different whiplash severities, we classified each county-month into mutually exclusive categories (no event, 5-year, 10-year, or 20-year event) and applied the corresponding severity-specific RR to estimate attributable deaths. Next, we examined the temporal trend in annual attributable deaths at the county level from 2003 to 2023 using the nonparametric Mann-Kendall test. We also aggregated the attributable burden separately for 2003–2012 and 2013–2023, annually by state and NOAA climate region, and nationally by year. Empirical 95% confidence intervals (CIs) were calculated as the 2.5th and 97.5th percentiles of 5000 Monte Carlo simulations of the estimated parameters from the main model.

We stratified the analysis by cause of death, specifically for infectious diseases (International Classification of Diseases Tenth Revision code: A00–B99), cardiovascular diseases (I00–I99), respiratory diseases (J00–J99), digestive diseases (K00–K95), endocrine and metabolic diseases (E00–E89), mental disorders (F01–F99), and external causes (V00–Y99). We also tested the potential effect modification by sex, age group (0–17, 18–64, and ≥65), education (secondary or lower and tertiary), race/ethnicity (non-Hispanic White, non-Hispanic Black, and Hispanic), household income (< median and ≥median), urbanicity (urban and rural), social vulnerability (<0.5 and ≥0.5), and Köppen-Geiger climate classification (dry, temperate, and continental). Tropical climate was excluded because few counties have this classification. We limited the subgroup analysis to the counties with a minimum population of 10,000 to ensure statistical power. We also stratified the analysis by transition pattern to test the potentially different associations with intrastate transitions and interstate changes.

We finally performed a series of sensitivity analyses to evaluate the robustness of our results. First, we changed the baseline period to 1961–1990 to recalculate whiplash thresholds. Second, we defined a whiplash event for a county when at least 50% of the population was exposed in a given month. Third, instead of identifying whiplash at the grid-cell level and then aggregating for each county, we first calculated population-weighted average SPEI for each county and then identified whiplash at the county level. Fourth, we conducted the analysis only in counties with a population of 25,000 or above. Fifth, we separately adjusted four model specifications in the first stage of analysis, including predefining a maximum lag of six months, estimating cumulative associations of 3 and 4 months, and setting three *df* for the lag-response association. Lastly, we included county-specific population in 2020 in the second stage of analysis.

All analyses were conducted in R software version 4.5.2. We used the packages *dlnm* and *mixmeta* for the regression analysis and *trend* for the trend test. This study was approved by the institutional review board at the University of Alabama at Birmingham and granted exemption as not human subject research.

## Results

Over 2003–2023, there were a total of 56,675,351 deaths recorded in the contiguous U.S. We observed considerable spatial heterogeneity in whiplash events (Figures 2, S1, and S2 for 5-, 10-, and 20-year whiplash respectively). Seasonal (SPEI-3) dry-to-wet events were more frequent in Florida, while wet-to-dry events were more common in southern New Mexico, Texas, and Louisiana. Most of California and southern Arizona experienced both directions of whiplash at seasonal and sub-annual scales. Across grid cells, we observed more seasonal than sub-annual whiplash events, and frequencies decreased with increasing RI (Table S1). The majority of consecutive-month SPEI changes that defined whiplash resulted in interstate transitions between wet and dry conditions rather than intrastate changes (i.e., intensification or recovery) within the same state (Table S2).

**Figure 2.**
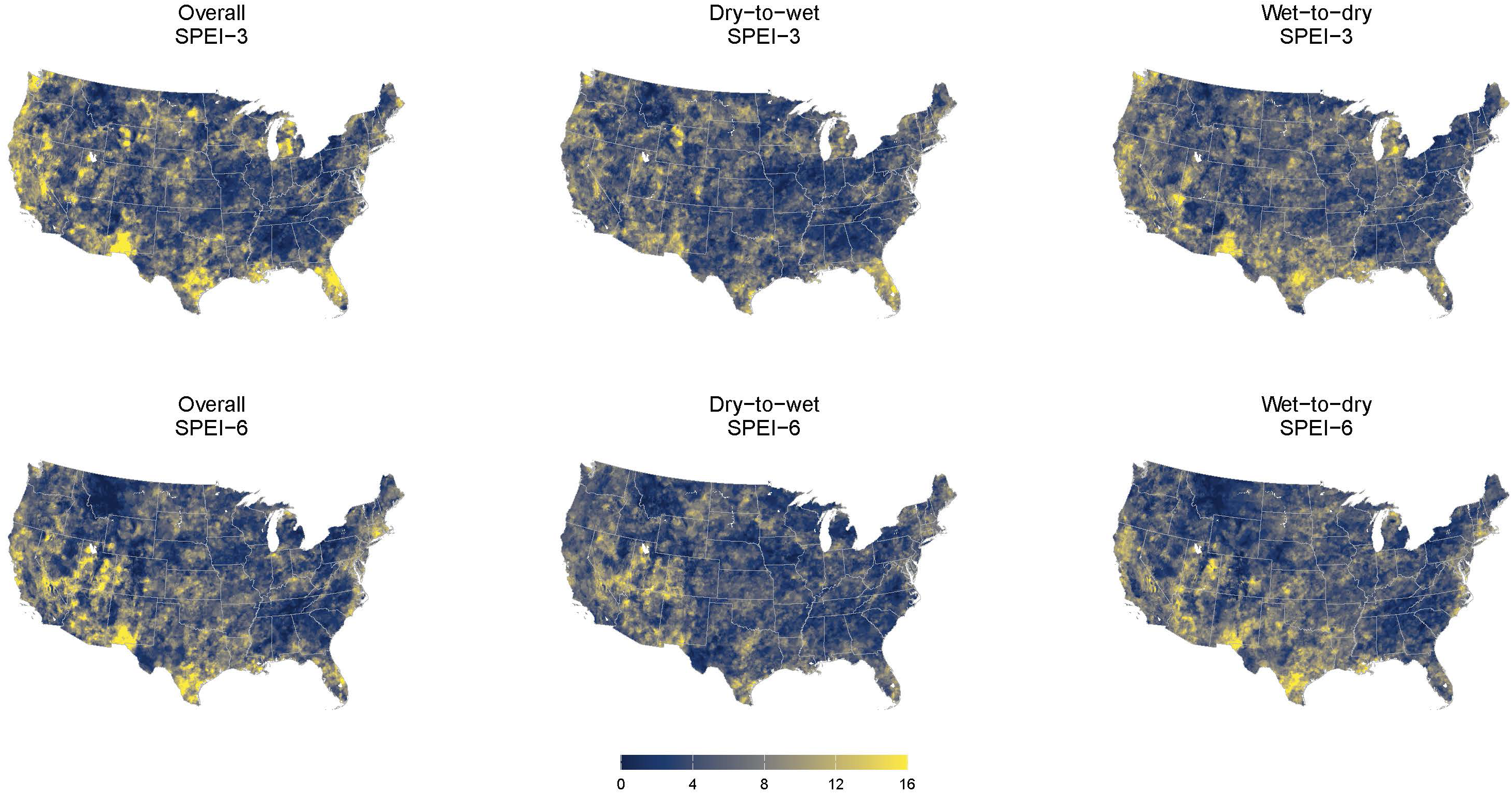
Frequencies of hydroclimate whiplash events with a 5-year recurrence interval during 2003–2023. The color scale is capped at the 99.5th percentile of the distribution of observed values to enhance visualization of spatial heterogeneity.

We observed a positive association between whiplash and all-cause mortality, with a generally shorter association lag for seasonal whiplash than for sub-annual whiplash (Figures 3A and S3). Over the 5-month cumulative lag, sub-annual whiplash showed consistently positive associations with mortality risk. We estimated similar RRs across event types under the same RI, while the RR increased with increasing RI under the same event type (Figure S3). Specifically, sub-annual overall whiplash with a 5-year RI increased the mortality risk by 3.4% (95% CI: 2.2–4.6), whereas seasonal overall whiplash showed a non-significant association with mortality (Figure 3B). For direction-specific whiplash events, wet-to-dry, but not dry-to-wet, seasonal whiplash significantly increased the mortality risk by 2.9% (95% CI: 1.8–4.1). In contrast, we observed significant association for both dry-to-wet and wet-to-dry sub-annual whiplash events (Figure 3B).

**Figure 3.**
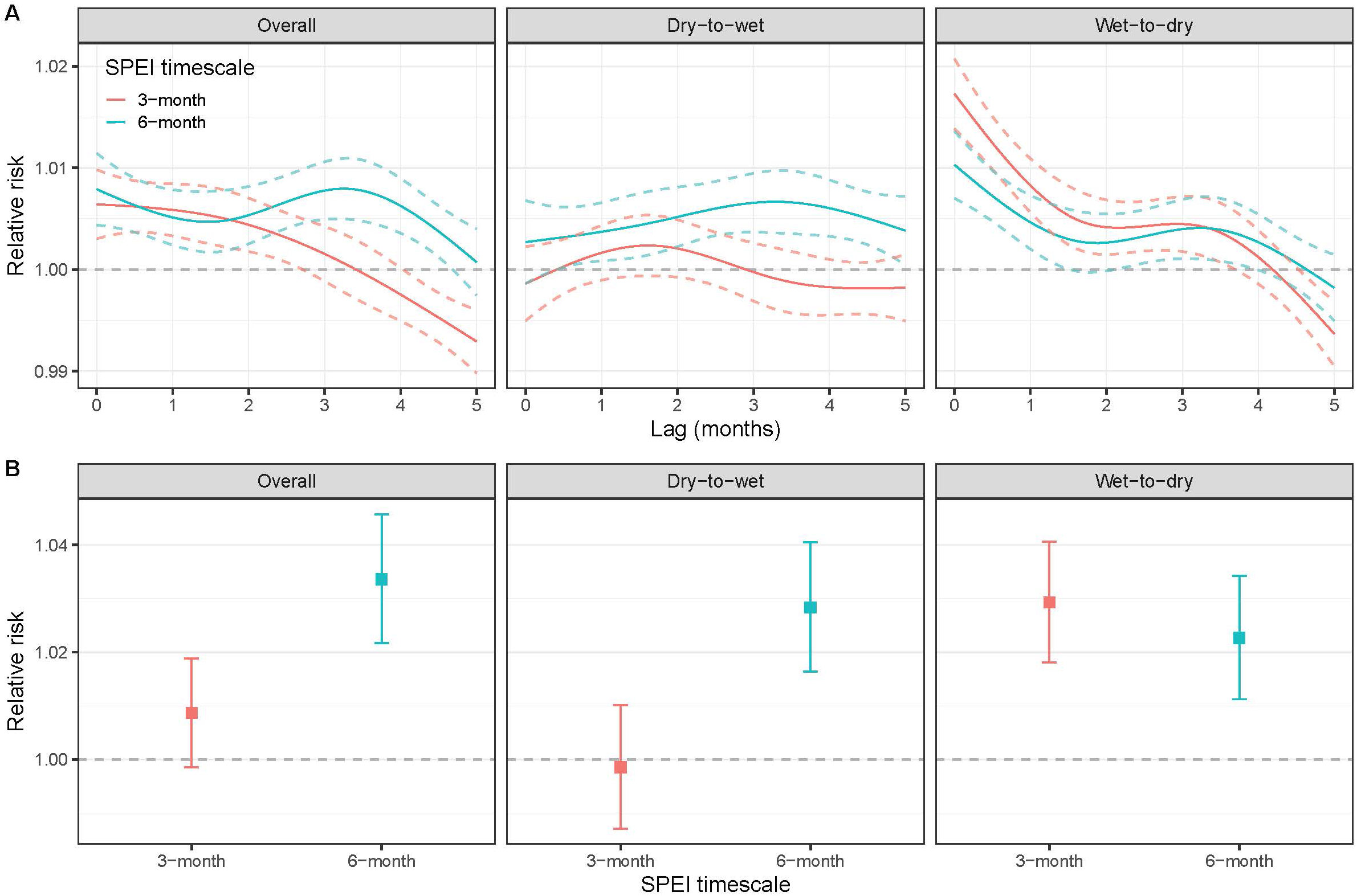
Lag-response association (A) and cumulative exposure-response association over lags of 0–5 months (B) between hydroclimate whiplash events with a 5-year recurrence interval and risk of all-cause mortality

From 2003 through 2023, a total of 103,471 (95% CI: 74,607–125,886; AF: 0.18%) deaths (or 4927 on average annually) were attributable to sub-annual overall whiplash, with the Northeast and Southeast showing the most attributable deaths and the Northwest and West showing the highest AF (Tables S3 and S4). When normalized by population, we observed higher annual attributable mortality in the West, South, and Great Plains (Figure 4). This attributable burden was driven by overall events for sub-annual whiplash but by wet-to-dry events for seasonal whiplash (Figures 4 and S4). During the study period, we estimated that almost three times as many deaths were attributable to sub-annual whiplash as to seasonal whiplash (AN: 34,899; AF: 0.06%). Furthermore, we found diverging trends in the mortality burden across all counties (Figure S5). Compared with 2003–2012, 1613 and 1478 counties showed increases and decreases in annual deaths attributable to sub-annual overall whiplash during 2013–2023, respectively, with more counties showing an increasing trend in the West. The overall trend test showed that from 2003 to 2023, 1521 counties exhibited a positive trend and 1524 exhibited a negative trend in overall-whiplash-attributable deaths, with more counties showing a significant increase in the Southwest and Kentucky, and more counties showing a significant decrease in the Northeast and Wisconsin (Figure S5B). Nationally, we did not observe a notable trend in the total number of deaths attributed to whiplash over time (Figures S6–S7). However, we observed an apparent increasing trend in mortality burden attributable to dry-to-wet whiplash in Florida, that attributable to wet-to-dry whiplash in the West (Washington, Oregon, California, Idaho, Utah, and Arizona) and Southeast (South Carolina, Alabama, and Georgia), and that attributable to overall whiplash in the West and Florida, particularly for sub-annual whiplash events (Figures 5, S8, and S9).

**Figure 4.**
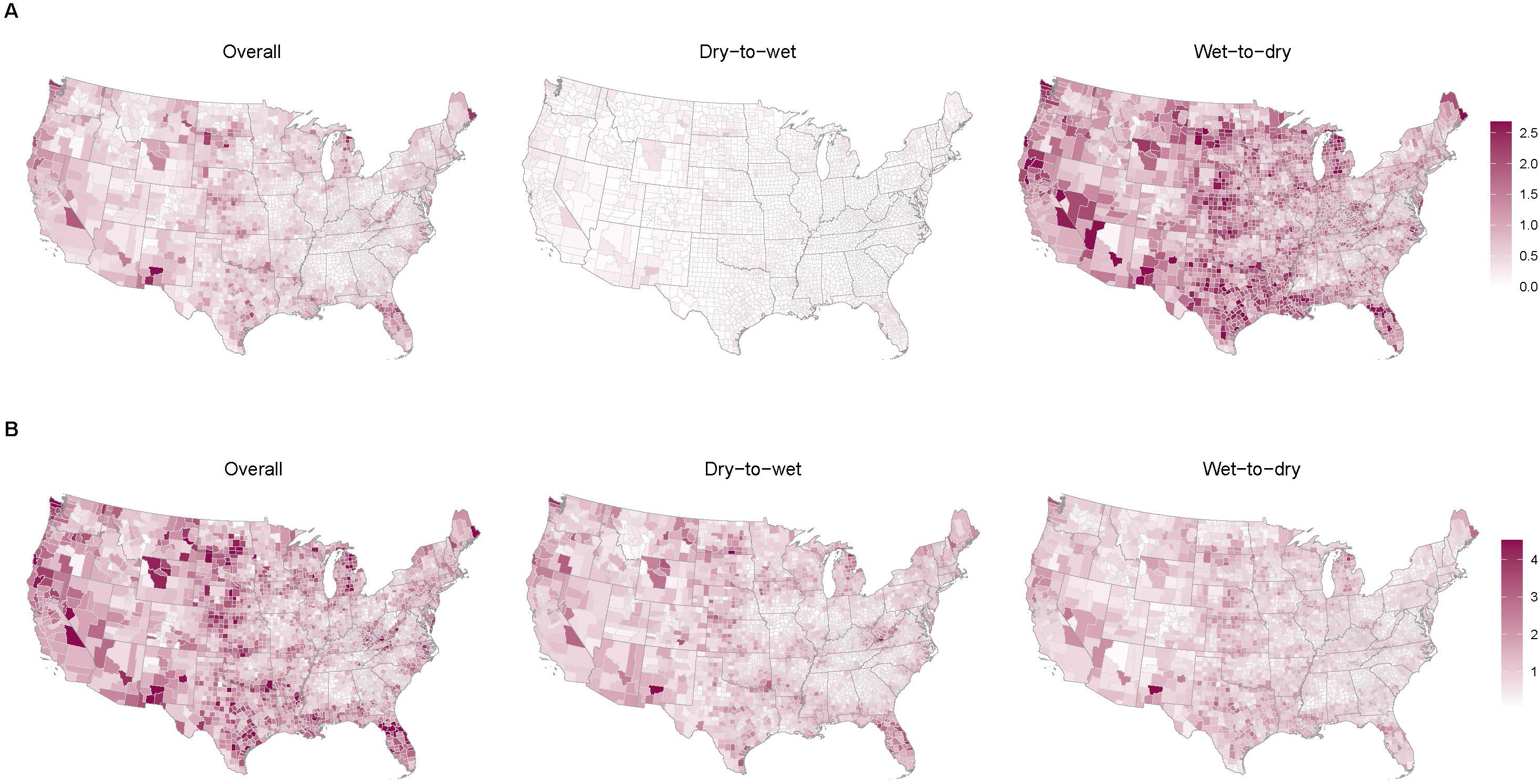
Average annual deaths attributable to seasonal (A) and sub-annual (B) hydroclimate whiplash events during 2003–2023 per 100,000 population. The color scale is bounded between the 0.5th and 99.5th percentiles of the distribution of average annual attributable deaths to enhance visualization of spatial heterogeneity.

**Figure 5.**
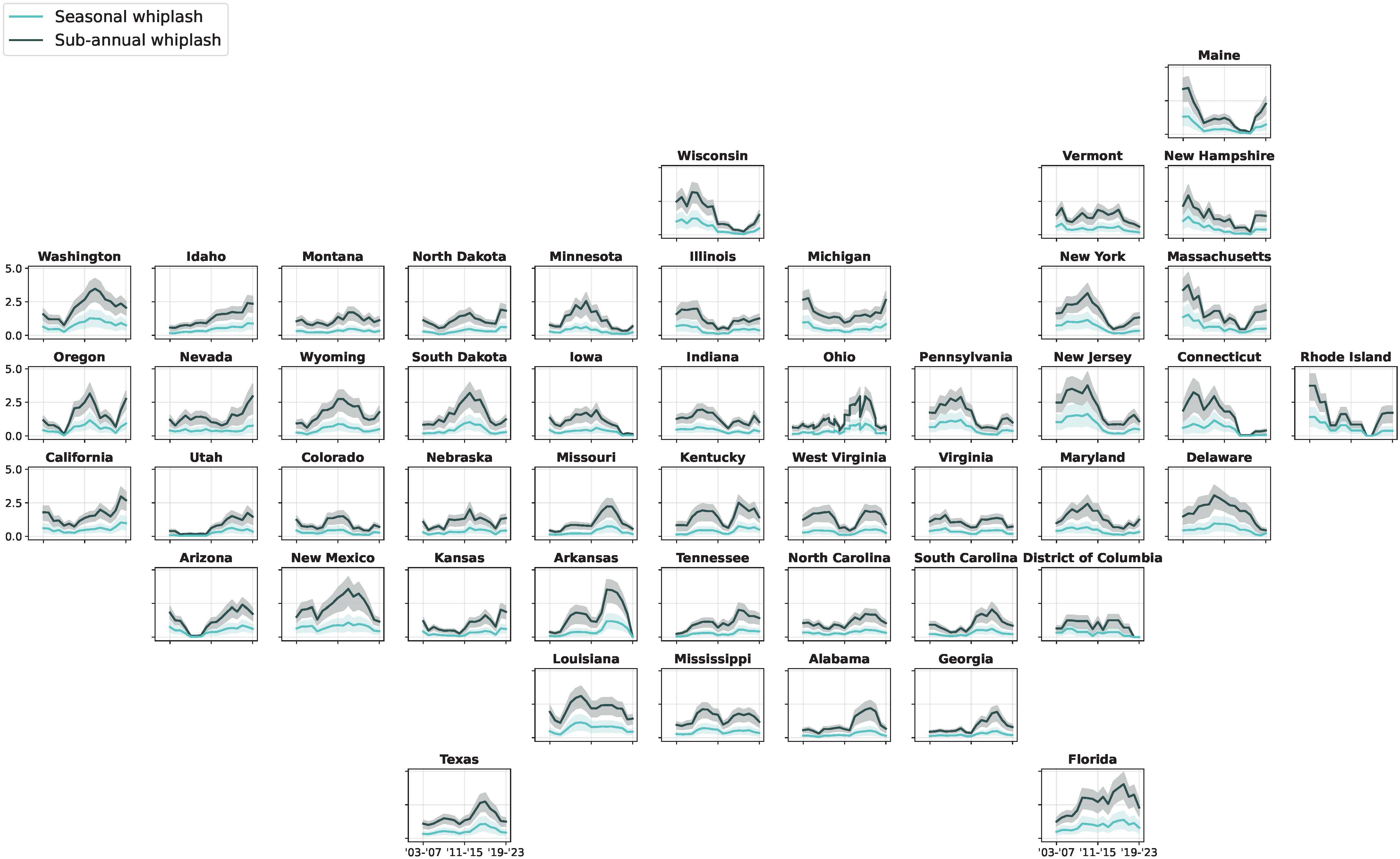
Five-year moving average annual number of deaths per 100,000 population attributable to overall hydroclimate whiplash events by state during 2003–2023. Shades represent the 95% confidence intervals, derived from 5000 Monte Carlo simulations of the estimated parameters from the main model.

We performed all stratified and sensitivity analyses using sub-annual whiplash with a 5-year RI to capture whiplash events across all severity levels. We found a positive association between whiplash and all types of cause-specific mortality, including infectious diseases, cardiovascular diseases, respiratory diseases, digestive diseases, endocrine diseases, mental disorders, and external causes over a lag of five months, with a notably stronger association for infectious diseases (RR: 1.45; 95% CI: 1.36–1.54) (Table 1). We observed a significantly stronger association across event types in younger populations (<65 years) than in the elderly (≥65 years) and a stronger association in the Hispanic population than in the non-Hispanic White and non-Hispanic Black populations (*p*<0.05 for the differences) (Figure S10). When stratified by transition pattern, estimates for overall and directional whiplash were similar across strata, with no clear evidence of effect modification (Table S5). We found only non-meaningful changes in the association estimates across all sensitivity analyses (Table S6).

**Table 1.** Cumulative associations between sub-annual hydroclimate whiplash events with a 5-year recurrence interval and risk of all-cause and cause-specific mortality over lags of 0–5 months.

|  | Overall whiplash | Dry-to-wet whiplash | Wet-to-dry whiplash |
| --- | --- | --- | --- |
| All-cause mortality | 1.03<br>(1.02, 1.05) | 1.03<br>(1.02, 1.04) | 1.02<br>(1.01, 1.03) |
| Infectious diseases | 1.45<br>(1.36, 1.54) | 1.45<br>(1.36, 1.55) | 1.44<br>(1.35, 1.52) |
| Circulatory diseases | 1.03<br>(1.02, 1.05) | 1.04<br>(1.02, 1.06) | 1.03<br>(1.02, 1.05) |
| Respiratory diseases | 1.09<br>(1.06, 1.12) | 1.11<br>(1.07, 1.14) | 1.07<br>(1.04, 1.10) |
| Digestive diseases | 1.17<br>(1.12, 1.23) | 1.18<br>(1.13, 1.24) | 1.17<br>(1.12, 1.22) |
| Endocrine diseases | 1.17<br>(1.12, 1.22) | 1.18<br>(1.13, 1.23) | 1.18<br>(1.13, 1.23) |
| Mental disorders | 1.23<br>(1.17, 1.29) | 1.22<br>(1.16, 1.28) | 1.19<br>(1.14, 1.25) |
| External causes | 1.11<br>(1.07, 1.15) | 1.07<br>(1.03, 1.11) | 1.10<br>(1.07, 1.14) |

## Discussion

This study employed a state-of-the-art and comprehensive definition of hydroclimate whiplash events and found a positive association between both overall and directional whiplash and mortality risk in the contiguous United States. A higher RR was estimated for whiplash with a longer RI representing events with lower frequency and higher severity. In addition, we found a significantly higher risk of infectious disease mortality associated with whiplash events. Nationally, we estimated that there were nearly 5000 annual deaths during 2003–2023 attributable to sub-annual whiplash.

Both drought and floods are associated with a higher risk of mortality.^11^ Although few epidemiologic studies have directly evaluated hydroclimate whiplash as an exposure, rapid transitions between dry and wet conditions can plausibly amplify health risks by combining the adverse consequences of drought, heavy precipitation, flooding, and post-event disruption, demonstrating multifactorial mechanisms linking whiplash to mortality. Dry-to-wet transitions may increase runoff over dry or compacted soils, overwhelm drainage and wastewater systems, mobilize pathogens and chemical contaminants into surface water, and create standing-water habitats favorable for vectors.^25,26^ Wet-to-dry transitions may contribute to water scarcity, heat stress, dust exposure, wildfire risk, degradation of water quality, and delayed recovery from preceding wet conditions.^27,28^ These pathways are consistent with broader evidence that floods and droughts are associated with infectious, respiratory, cardiovascular, injury-related, and other health outcomes,^29,30^ even though the specific epidemiologic consequences of sequential hydroclimate transitions remain under-characterized.

The observed stronger association with infectious disease mortality may reflect several plausible, though unconfirmed, pathways. Dry-to-wet transitions can increase exposure to waterborne pathogens when heavy rainfall follows drought-hardened soils, and may also promote fungal growth or alter vector habitats and host-vector dynamics. Such hydroclimatic swings have been linked to diarrhea, Valley Fever, and mosquito-borne infections.^15–17^ Wet-to-dry transitions may likewise increase risk by concentrating contaminants in shrinking water sources, resuspending microbial or fungal particles in dust, and limiting hygiene through water scarcity. However, because infectious disease deaths encompass heterogeneous causes, these mechanisms require further epidemiological investigation.

Anthropogenic global warming increases the likelihood of not only individual hazards but also compound and cascading climate extremes.^31^ A study showed that compound drought-flood events caused eight times as many affected people and economic losses as isolated drought or flood events.^32^ We used the indicator of SPEI to characterize such dynamics. Although continuous SPEI does not capture the full spectrum of discrete drought and flood events, it reflects both precipitation deficits and atmospheric evaporative demand and therefore provides a physically consistent measure of hydroclimate anomalies across timescales.^22^ This multiscale perspective enables simultaneous assessment of transient and prolonged hydrological shifts, making SPEI an appropriate diagnostic metric for studying whiplash phenomena.

Whiplash events differ from other compound hazards in that they involve multiple events occurring in sequence over relatively short transition times, with evidence that these transition times are shortening across nearly 60% of the global land area under a warming climate.^6^ Indeed, with the increasing likelihood of flash droughts^33^ and flash floods^34^ under climate change, we can expect a higher frequency of rapid transitions between these states.^7^ The potential detrimental impacts of hydroclimate whiplash likely arise not only from the individual and combined physical hazards but also from rapid shifts in hydroclimatic conditions—whether transitions between states or abrupt changes within the same state—which can outpace the population’s adaptive capacity already compromised by prior exposures.^21^ In this study, we used only the difference between SPEI values for two adjacent months to identify whiplash, with the explicit purpose of recognizing rapid month-to-month changes in extreme states. However, we identified a lagged association with all-cause mortality, indicating a persistent health impact. Some indirect mechanisms might explain this prolonged association. For example, A study revealed a twofold risk of rice yield loss due to compound dry-and-wet extremes compared to isolated wet and dry extremes, with a 43% higher risk from dry-to-wet extremes than from wet-to-dry extremes.^35^ Future studies are needed to explore the long-term effects of whiplash events over multi-month timescales.

In our secondary analysis, we distinguished transitions between hydroclimate states from rapid intensification or recovery within the same state. Although both patterns reflect abrupt hydroclimate change, transitions between states may represent qualitatively different exposures from intrastate amplification or alleviation of existing conditions and may have distinct environmental and societal consequences. However, we observed largely consistent point estimates across strata, suggesting that the magnitude of abrupt hydroclimate change, rather than the distinction between intrastate intensification or recovery and interstate transitions, may be an important risk factor for mortality. These findings further indicate that the whiplash metric used in the main analysis provided a reasonable balance between statistical power and estimation stability. Future studies are needed to further evaluate whether transitions between hydroclimate states amplify risks of other health outcomes beyond those associated with the magnitude of change alone.

We acknowledge several limitations. First, the whiplash identification is sensitive to the choice of indicator, timescale, baseline period, whiplash event type, and RI, and the existing literature exhibits substantial methodological heterogeneity.^8^ Although alternative definitions may yield distinct exposure estimates, we addressed this subjectivity by examining multiple timescales, event types, and RIs. We also adopted a different baseline period and found robust associations. Second, the population-weighting approach used to upscale the binary exposure from the grid-cell to the county level is inherently subjective. Applying a higher threshold in the sensitivity analysis captured events affecting a larger population but reduced event frequency, highlighting a trade-off between meaningful representativeness and statistical power. The alternative approach based on population-weighted continuous SPEI is also imperfect: it avoids subjective thresholds for county-level event identification but may obscure localized signals at the grid-cell level. Third, we used nested thresholds for whiplash definitions to examine the association, which may bias mortality burden estimates. Although multi-level whiplash indicators with mutually exclusive categories would be conceptually preferable, such models showed convergence failure and unstable variance estimates in the second-stage meta-analysis, likely due to the rarity of events. Our estimates, therefore, reflect the risk associated with events exceeding a given severity threshold; applying them to mutually exclusive categories approximates a stepwise exposure–response relationship across increasing severity levels.

In conclusion, this nationwide study shows that hydroclimate whiplash was associated with increased mortality risk, underscoring its public health relevance of rapid transitions between wet and dry conditions. As the frequency of these events is projected to rise under climate change,^7^ existing health protection and adaptation strategies, which are largely designed for isolated hazards, may be insufficient. Our findings support the need for integrated approaches that explicitly address sequential and compound extremes, including strengthened early warning systems and coordinated preparedness across water, food, and healthcare systems. Prioritizing populations most vulnerable to these rapid transitions will be essential to reducing preventable mortality and enhancing climate resilience.

## Acknowledgement

A.M.A. and P.W. conceived and designed the study and prepared and cleaned the data. P.W. developed the analytical plan, performed the exposure assessment and statistical analysis, and wrote the original draft of the manuscript. A.M.A., Y.M., and J.D.S. contributed to the interpretation of the results, critically reviewed the manuscript, and edited the original version. All authors have approved the final draft of the manuscript. We declare no competing interests. The authors received no funding for this work.

## Data sharing statement

Mortality data were obtained from the National Center for Health Statistics Multiple Cause of Death restricted-use files, which are not publicly available due to confidentiality protections and require an approved data use agreement. The standardized precipitation evapotranspiration index at a resolution of 0.05° (∼ 5 km × 5 km) is publicly available from the National Oceanic and Atmospheric Administration’s National Centers for Environmental Information at https://www.drought.gov/data-maps-tools/us-gridded-standardized-precipitation-index-spei-nclimgrid-monthly.

## Code availability

The R programming code for the main analysis is publicly available at https://github.com/pwumd/whiplash.

