## Supplementary information for "Mortality risk and burden associated with hydroclimate whiplash in the United States"

Pin Wang, Yiqun Ma, Jennifer D. Stowell, Azar M. Abadi

**Text S1 Supplementary method**

In this study, we focused on whiplash events defined using the standardized precipitation evapotranspiration index (SPEI) at 3- and 6-month accumulation periods, as these timescales provide a balance between physical realism and epidemiological interpretability. Shorter timescales (e.g., SPEI-1) are highly sensitive to transient monthly variability and may capture short-lived fluctuations that are less relevant to sustained hydroclimatic stress, leading to misclassification of exposure and unstable or counterintuitive associations with health outcomes. In contrast, longer timescales (e.g., SPEI-12) reflect hydroclimatic conditions that evolve gradually over time. Therefore, imposing a definition based on rapid month-to-month transitions may be inconsistent with the underlying physical dynamics of drought development, thereby making these events inherently rare and reducing statistical power to detect associations. SPEI-3 and SPEI-6, therefore, represent intermediate scales that better capture meaningful and epidemiologically relevant hydroclimatic transitions.

**Table S1** Descriptive statistics for the total counts of hydroclimate whiplash events during 2003–2023 across all grid cells in the contiguous United States by timescale, recurrence interval, and event type

|  | Mean (SD) | Min. | Median | Max. | IQR |
| --- | --- | --- | --- | --- | --- |
| Seasonal (3-month timescale) |  |  |  |  |  |
| 5-year interval |  |  |  |  |  |
| Overall | 6.3 (3.6) | 0 | 6.0 | 31.0 | 4.0–8.0 |
| Dry-to-wet | 5.6 (2.9) | 0 | 5.0 | 22.0 | 4.0–7.0 |
| Wet-to-dry | 6.0 (2.9) | 0 | 6.0 | 25.0 | 4.0–8.0 |
| 10-year interval |  |  |  |  |  |
| Overall | 3.6 (2.7) | 0 | 3.0 | 24.0 | 2.0–5.0 |
| Dry-to-wet | 3.2 (2.2) | 0 | 3.0 | 17.0 | 2.0–4.0 |
| Wet-to-dry | 3.4 (2.2) | 0 | 3.0 | 21.0 | 2.0–5.0 |
| 20-year interval |  |  |  |  |  |
| Overall | 2.2 (2.1) | 0 | 2.0 | 20.0 | 1.0–3.0 |
| Dry-to-wet | 1.9 (1.7) | 0 | 2.0 | 15.0 | 1.0–3.0 |
| Wet-to-dry | 2.0 (1.8) | 0 | 2.0 | 15.0 | 1.0–3.0 |
| Sub-annual (6-month timescale) |  |  |  |  |  |
| 5-year interval |  |  |  |  |  |
| Overall | 5.9 (3.4) | 0 | 5.0 | 27.0 | 3.0–8.0 |
| Dry-to-wet | 5.2 (2.7) | 0 | 5.0 | 22.0 | 3.0–7.0 |
| Wet-to-dry | 5.7 (3.0) | 0 | 5.0 | 22.0 | 4.0–7.0 |
| 10-year interval |  |  |  |  |  |
| Overall | 3.5 (2.6) | 0 | 3.0 | 23.0 | 2.0–5.0 |
| Dry-to-wet | 2.9 (2.0) | 0 | 3.0 | 15.0 | 1.0–4.0 |
| Wet-to-dry | 3.2 (2.2) | 0 | 3.0 | 17.0 | 2.0–4.0 |
| 20-year interval |  |  |  |  |  |
| Overall | 2.2 (2.0) | 0 | 2.0 | 21.0 | 1.0–3.0 |
| Dry-to-wet | 1.8 (1.6) | 0 | 2.0 | 14.0 | 1.0–3.0 |
| Wet-to-dry | 2.0 (1.7) | 0 | 2.0 | 15.0 | 1.0–3.0 |

Note: SD: standard deviation; IQR: interquartile range

**Table S2** Mean and standard deviation of the total counts of hydroclimate whiplash events during 2003–2023 across all grid cells in the contiguous United States by timescale, recurrence interval, event type, and transition pattern

|  | Seasonal (3-month timescale) | | Sub-annual (6-month timescale) | |
| --- | --- | --- | --- | --- |
|  | Interstate transition | Intrastate change | Interstate transition | Intrastate change |
| 5-year interval |  |  |  |  |
| Overall | 4.7 (2.8) | 1.6 (1.6) | 3.5 (2.2) | 2.3 (2.0) |
| Dry-to-wet | 3.9 (2.2) | 1.7 (1.5) | 2.8 (1.7) | 2.4 (1.9) |
| Wet-to-dry | 4.3 (2.2) | 1.9 (1.6) | 3.1 (1.8) | 2.6 (1.9) |
| 10-year interval |  |  |  |  |
| Overall | 2.8 (2.2) | 0.8 (1.1) | 2.3 (1.8) | 1.2 (1.4) |
| Dry-to-wet | 2.4 (1.8) | 0.8 (1.0) | 1.7 (1.4) | 1.2 (1.3) |
| Wet-to-dry | 2.6 (1.7) | 0.9 (1.1) | 1.9 (1.5) | 1.3 (1.3) |
| 20-year interval |  |  |  |  |
| Overall | 1.8 (1.8) | 0.4 (0.8) | 1.5 (1.5) | 0.7 (1.0) |
| Dry-to-wet | 1.5 (1.4) | 0.4 (0.7) | 1.2 (1.2) | 0.7 (0.9) |
| Wet-to-dry | 1.6 (1.4) | 0.5 (0.8) | 1.3 (1.2) | 0.7 (1.0) |

Note: An interstate transition represents a regime shift between the wet and dry states; an intrastate change represents intensification or recovery within the wet or dry state.

**Table S3** Annual attributable burden of all-cause mortality associated with sub-annual hydroclimate whiplash events by state

| State | Overall whiplash | | Dry-to-wet whiplash | | Wet-to-dry whiplash | |
| --- | --- | --- | --- | --- | --- | --- |
|  | Deaths | Fraction (%) | Deaths | Fraction (%) | Deaths | Fraction (%) |
| Alabama | 46  (29, 57) | 0.09  (0.05, 0.11) | 33  (25, 41) | 0.06  (0.05, 0.08) | 31  (22, 40) | 0.06  (0.04, 0.08) |
| Arizona | 98  (74, 119) | 0.18  (0.13, 0.22) | 77  (59, 96) | 0.14  (0.11, 0.17) | 57  (40, 73) | 0.10  (0.07, 0.13) |
| Arkansas | 42  (29, 51) | 0.13  (0.09, 0.16) | 23  (18, 29) | 0.07  (0.06, 0.09) | 24  (17, 30) | 0.08  (0.05, 0.10) |
| California | 697  (505, 846) | 0.27  (0.19, 0.33) | 344  (255, 429) | 0.13  (0.10, 0.17) | 342  (242, 438) | 0.13  (0.09, 0.17) |
| Colorado | 55  (39, 67) | 0.15  (0.11, 0.19) | 50  (39, 62) | 0.14  (0.11, 0.17) | 29  (20, 37) | 0.08  (0.06, 0.10) |
| Connecticut | 51  (35, 63) | 0.17  (0.11, 0.21) | 29  (22, 36) | 0.10  (0.07, 0.12) | 27  (19, 35) | 0.09  (0.06, 0.11) |
| Delaware | 17  (12, 20) | 0.19  (0.13, 0.24) | 10  (7, 12) | 0.11  (0.08, 0.14) | 8  (6, 10) | 0.10  (0.07, 0.12) |
| District of Columbia | 5  (4, 7) | 0.10  (0.07, 0.13) | 5  (4, 7) | 0.10  (0.07, 0.13) | 2  (1, 3) | 0.04  (0.03, 0.06) |
| Florida | 538  (398, 651) | 0.28  (0.21, 0.34) | 393  (295, 488) | 0.20  (0.15, 0.25) | 207  (146, 267) | 0.11  (0.08, 0.14) |
| Georgia | 81  (55, 99) | 0.10  (0.07, 0.12) | 43  (33, 53) | 0.05  (0.04, 0.07) | 46  (33, 58) | 0.06  (0.04, 0.07) |
| Idaho | 24  (17, 29) | 0.18  (0.13, 0.22) | 15  (11, 18) | 0.12  (0.09, 0.14) | 10  (7, 13) | 0.08  (0.05, 0.10) |
| Illinois | 156  (112, 189) | 0.14  (0.10, 0.17) | 67  (51, 82) | 0.06  (0.05, 0.08) | 109  (78, 139) | 0.10  (0.07, 0.13) |
| Indiana | 85  (61, 103) | 0.14  (0.10, 0.17) | 37  (28, 45) | 0.06  (0.05, 0.07) | 60  (42, 76) | 0.10  (0.07, 0.12) |
| Iowa | 33  (23, 41) | 0.11  (0.08, 0.14) | 21  (16, 25) | 0.07  (0.05, 0.09) | 20  (15, 26) | 0.07  (0.05, 0.09) |
| Kansas | 34  (24, 41) | 0.13  (0.09, 0.15) | 20  (16, 25) | 0.08  (0.06, 0.09) | 23  (16, 30) | 0.09  (0.06, 0.11) |
| Kentucky | 59  (38, 73) | 0.13  (0.08, 0.16) | 33  (25, 40) | 0.07  (0.05, 0.09) | 39  (28, 49) | 0.08  (0.06, 0.11) |
| Louisiana | 100  (71, 121) | 0.22  (0.16, 0.27) | 44  (34, 54) | 0.10  (0.08, 0.12) | 62  (44, 81) | 0.14  (0.10, 0.18) |
| Maine | 23  (17, 28) | 0.17  (0.12, 0.20) | 21  (16, 26) | 0.15  (0.11, 0.19) | 9  (6, 11) | 0.06  (0.04, 0.08) |
| Maryland | 80  (54, 98) | 0.17  (0.11, 0.20) | 57  (42, 70) | 0.12  (0.09, 0.15) | 35  (25, 44) | 0.07  (0.05, 0.09) |
| Massachusetts | 129  (95, 158) | 0.23  (0.17, 0.28) | 93  (69, 117) | 0.16  (0.12, 0.21) | 52  (37, 67) | 0.09  (0.06, 0.12) |
| Michigan | 199  (140, 242) | 0.21  (0.15, 0.25) | 119  (90, 146) | 0.13  (0.09, 0.15) | 117  (84, 148) | 0.12  (0.09, 0.16) |
| Minnesota | 66  (45, 81) | 0.16  (0.11, 0.19) | 44  (33, 54) | 0.10  (0.08, 0.13) | 41  (29, 52) | 0.10  (0.07, 0.12) |
| Mississippi | 42  (30, 51) | 0.13  (0.09, 0.16) | 31  (24, 38) | 0.10  (0.07, 0.12) | 23  (16, 29) | 0.07  (0.05, 0.09) |
| Missouri | 60  (42, 72) | 0.10  (0.07, 0.12) | 37  (28, 45) | 0.06  (0.05, 0.08) | 40  (29, 50) | 0.07  (0.05, 0.08) |
| Montana | 12  (8, 15) | 0.13  (0.08, 0.15) | 6  (5, 8) | 0.06  (0.05, 0.08) | 6  (5, 8) | 0.07  (0.05, 0.09) |
| Nebraska | 24  (16, 29) | 0.14  (0.10, 0.18) | 9  (7, 12) | 0.06  (0.04, 0.07) | 14  (10, 18) | 0.09  (0.06, 0.11) |
| Nevada | 48  (33, 59) | 0.21  (0.14, 0.26) | 22  (17, 27) | 0.10  (0.07, 0.12) | 28  (20, 36) | 0.12  (0.09, 0.16) |
| New Hampshire | 20  (15, 24) | 0.17  (0.13, 0.21) | 14  (11, 17) | 0.12  (0.09, 0.15) | 8  (5, 11) | 0.07  (0.05, 0.09) |
| New Jersey | 173  (130, 214) | 0.23  (0.18, 0.29) | 101  (74, 127) | 0.14  (0.10, 0.17) | 66  (42, 88) | 0.09  (0.06, 0.12) |
| New Mexico | 43  (30, 52) | 0.24  (0.17, 0.29) | 22  (17, 28) | 0.12  (0.09, 0.15) | 24  (17, 31) | 0.13  (0.10, 0.17) |
| New York | 304  (227, 369) | 0.19  (0.14, 0.24) | 209  (153, 263) | 0.13  (0.10, 0.17) | 117  (80, 152) | 0.07  (0.05, 0.10) |
| North Carolina | 112  (78, 136) | 0.13  (0.09, 0.16) | 66  (50, 82) | 0.08  (0.06, 0.09) | 56  (40, 71) | 0.06  (0.05, 0.08) |
| North Dakota | 10  (6, 12) | 0.15  (0.10, 0.19) | 7  (5, 8) | 0.10  (0.08, 0.13) | 4  (3, 5) | 0.07  (0.05, 0.08) |
| Ohio | 129  (89, 158) | 0.11  (0.08, 0.13) | 80  (61, 100) | 0.07  (0.05, 0.08) | 77  (55, 97) | 0.06  (0.05, 0.08) |
| Oklahoma | 48  (33, 59) | 0.12  (0.08, 0.15) | 29  (22, 36) | 0.07  (0.06, 0.09) | 23  (17, 30) | 0.06  (0.04, 0.08) |
| Oregon | 74  (54, 90) | 0.21  (0.15, 0.25) | 47  (36, 58) | 0.13  (0.10, 0.17) | 39  (28, 49) | 0.11  (0.08, 0.14) |
| Pennsylvania | 191  (144, 232) | 0.14  (0.11, 0.17) | 114  (87, 141) | 0.09  (0.07, 0.11) | 95  (65, 125) | 0.07  (0.05, 0.09) |
| Rhode Island | 18  (13, 22) | 0.18  (0.13, 0.22) | 15  (11, 18) | 0.15  (0.11, 0.18) | 7  (5, 9) | 0.07  (0.05, 0.09) |
| South Carolina | 50  (34, 62) | 0.11  (0.07, 0.13) | 34  (26, 41) | 0.07  (0.06, 0.09) | 30  (21, 38) | 0.06  (0.05, 0.08) |
| South Dakota | 14  (9, 18) | 0.18  (0.12, 0.23) | 7  (5, 8) | 0.09  (0.07, 0.11) | 7  (5, 9) | 0.09  (0.07, 0.12) |
| Tennessee | 67  (43, 83) | 0.10  (0.06, 0.13) | 38  (29, 48) | 0.06  (0.04, 0.07) | 54  (39, 69) | 0.08  (0.06, 0.10) |
| Texas | 451  (335, 546) | 0.24  (0.18, 0.29) | 253  (191, 312) | 0.13  (0.10, 0.17) | 262  (181, 341) | 0.14  (0.10, 0.18) |
| Utah | 26  (18, 32) | 0.15  (0.11, 0.19) | 22  (16, 28) | 0.13  (0.10, 0.17) | 16  (11, 20) | 0.09  (0.07, 0.12) |
| Vermont | 8  (6, 10) | 0.14  (0.11, 0.17) | 7  (5, 9) | 0.13  (0.09, 0.16) | 3  (2, 4) | 0.05  (0.04, 0.07) |
| Virginia | 89  (63, 109) | 0.14  (0.10, 0.17) | 61  (45, 76) | 0.09  (0.07, 0.11) | 41  (29, 52) | 0.06  (0.04, 0.08) |
| Washington | 164  (125, 198) | 0.31  (0.23, 0.37) | 85  (63, 106) | 0.16  (0.12, 0.20) | 83  (55, 109) | 0.15  (0.10, 0.20) |
| West Virginia | 23  (14, 28) | 0.10  (0.06, 0.12) | 21  (16, 26) | 0.09  (0.07, 0.11) | 11  (8, 14) | 0.05  (0.03, 0.06) |
| Wisconsin | 99  (73, 120) | 0.19  (0.14, 0.24) | 60  (45, 75) | 0.12  (0.09, 0.15) | 46  (32, 59) | 0.09  (0.06, 0.12) |
| Wyoming | 10  (7, 12) | 0.20  (0.14, 0.25) | 6  (4, 7) | 0.12  (0.09, 0.15) | 5  (3, 6) | 0.10  (0.07, 0.12) |

Note: Numbers in parentheses indicate 95% confidence intervals derived from 5000 Monte Carlo simulations of the estimated parameters from the main model.

**Table S4** Annual attributable burden of all-cause mortality associated with sub-annual hydroclimate whiplash events by climate region defined by the National Oceanic and Atmospheric Administration

| Climate region | Overall whiplash | | Dry-to-wet whiplash | | Wet-to-dry whiplash | |
| --- | --- | --- | --- | --- | --- | --- |
|  | Deaths | Fraction (%) | Deaths | Fraction (%) | Deaths | Fraction (%) |
| Northeast | 998  (741, 1214) | 0.18  (0.14, 0.22) | 660  (491, 822) | 0.12  (0.09, 0.15) | 417  (290, 541) | 0.08  (0.05, 0.10) |
| Northern Rockies and Plains | 69  (47, 85) | 0.15  (0.10, 0.19) | 35  (27, 43) | 0.08  (0.06, 0.10) | 37  (26, 47) | 0.08  (0.06, 0.10) |
| Northwest | 283  (212, 344) | 0.25  (0.18, 0.30) | 161  (120, 200) | 0.14  (0.10, 0.17) | 142  (98, 184) | 0.12  (0.09, 0.16) |
| Ohio Valley | 579  (402, 706) | 0.12  (0.08, 0.15) | 313  (241, 385) | 0.06  (0.05, 0.08) | 389  (279, 494) | 0.08  (0.06, 0.10) |
| South | 717  (521, 869) | 0.20  (0.14, 0.24) | 400  (304, 493) | 0.11  (0.08, 0.14) | 418  (292, 539) | 0.12  (0.08, 0.15) |
| Southeast | 916  (658, 1112) | 0.17  (0.13, 0.21) | 630  (476, 780) | 0.12  (0.09, 0.15) | 410  (294, 523) | 0.08  (0.06, 0.10) |
| Southwest | 222  (161, 270) | 0.18  (0.13, 0.21) | 172  (130, 213) | 0.14  (0.10, 0.17) | 125  (89, 160) | 0.10  (0.07, 0.13) |
| Upper Midwest | 397  (282, 482) | 0.18  (0.13, 0.22) | 243  (185, 300) | 0.11  (0.08, 0.14) | 224  (161, 286) | 0.10  (0.07, 0.13) |
| West | 745  (538, 905) | 0.26  (0.19, 0.32) | 365  (272, 456) | 0.13  (0.10, 0.16) | 370  (263, 473) | 0.13  (0.09, 0.17) |

Note: Numbers in parentheses indicate 95% confidence intervals derived from 5000 Monte Carlo simulations of the estimated parameters from the main model. Northeast region includes the states of Connecticut, Delaware, Maine, Maryland, Massachusetts, New Hampshire, New Jersey, New York, Pennsylvania, Rhode Island, and Vermont; Northern Rockies and Plains region includes the states of Montana, Nebraska, North Dakota, South Dakota, and Wyoming; Northwest region includes the states of Idaho, Oregon, and Washington; Ohio Valley includes the states of Illinois, Indiana, Kentucky, Missouri, Ohio, Tennessee, and West Virginia; South region includes the states of Arkansas, Kansas, Louisiana, Mississippi, Oklahoma, and Texas; Southeast region includes the states of Alabama, Florida, Georgia, North Carolina, South Carolina, and Virginia; Southwest region includes the states of Arizona, Colorado, New Mexico, and Utah; Upper Midwest region includes the states of Iowa, Michigan, Minnesota, and Wisconsin; West region includes the states of California and Nevada.

**Table S5** Stratified analysis of the association between all-cause mortality and sub-annual hydroclimate whiplash events with a 5-year recurrence interval, by transition pattern

|  | Overall whiplash | Dry-to-wet whiplash | Wet-to-dry whiplash |
| --- | --- | --- | --- |
| Main analysis | 1.03  (1.02, 1.05) | 1.03  (1.02, 1.04) | 1.02  (1.01, 1.03) |
| Interstate transition | 1.04  (1.02, 1.05) | 1.04  (1.03, 1.06) | 1.03  (1.01, 1.05) |
| Intrastate change | 1.04  (1.02, 1.06) | 1.03  (1.01, 1.05) | 1.03  (1.01, 1.05) |

Note: An interstate transition represents a regime shift between the wet and dry states; an intrastate transition represents intensification or recovery within the wet or dry state.

**Table S6** Sensitivity analyses for the association between all-cause mortality and sub-annual hydroclimate whiplash events with a 5-year recurrence interval

|  | Overall whiplash | Dry-to-wet whiplash | Wet-to-dry whiplash |
| --- | --- | --- | --- |
| Main analysis | 1.03  (1.02, 1.05) | 1.03  (1.02, 1.04) | 1.02  (1.01, 1.03) |
| Baseline period of 1961-1990 | 1.03  (1.02, 1.05) | 1.03  (1.02, 1.04) | 1.03  (1.02, 1.04) |
| Weighted average threshold of 0.5 | 1.04  (1.02, 1.05) | 1.04  (1.03, 1.05) | 1.02  (1.01, 1.03) |
| Weighted average SPEI by county | 1.04  (1.03, 1.05) | 1.04  (1.03, 1.05) | 1.02  (1.01, 1.04) |
| Counties with a population over 25,000 | 1.03  (1.02, 1.04) | 1.02  (1.01, 1.04) | 1.02  (1.01, 1.03) |
| DLNM – maximum lag of 6 months | 1.04  (1.02, 1.05) | 1.03  (1.02, 1.04) | 1.02  (1.01, 1.03) |
| DLNM – cumulative effect of 3 months | 1.03  (1.02, 1.04) | 1.02  (1.01, 1.03) | 1.02  (1.01, 1.03) |
| DLNM – cumulative effect of 4 months | 1.03  (1.02, 1.04) | 1.02  (1.01, 1.04) | 1.02  (1.01, 1.03) |
| DLNM – 3 degrees of freedom for lag | 1.03  (1.02, 1.04) | 1.02  (1.01, 1.04) | 1.02  (1.01, 1.03) |
| Population as meta-predictor | 1.03  (1.02, 1.05) | 1.03  (1.02, 1.04) | 1.02  (1.01, 1.03) |

Note: SPEI: standardized precipitation evapotranspiration index. DLNM: distributed lag non-linear model.

**Figure S1** Frequencies of hydroclimate whiplash events with a 10-year recurrence interval during 2003–2023. The color scale is capped at the 99.5th percentile of the distribution of observed values to enhance visualization of spatial heterogeneity.

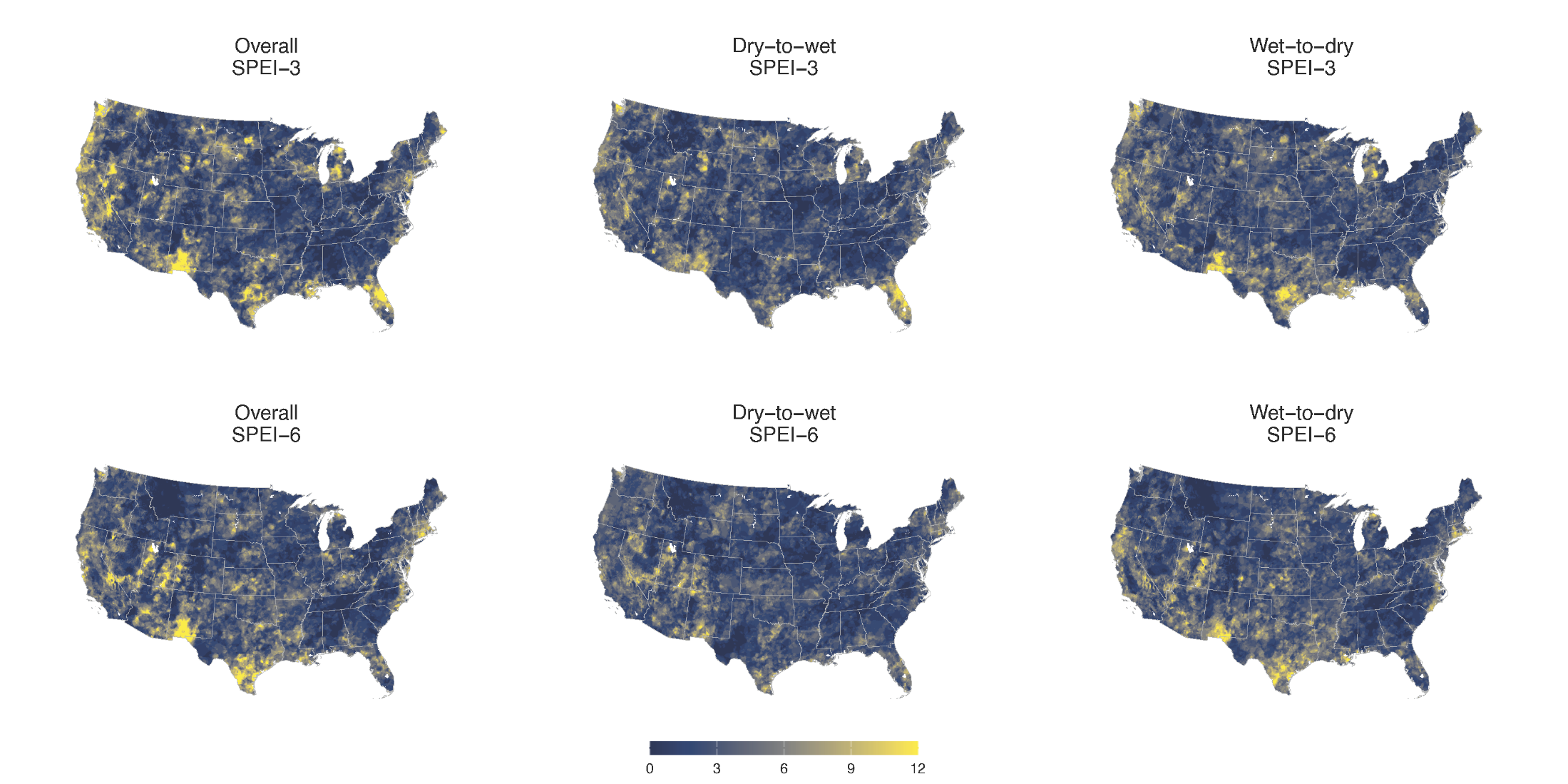

**Figure S2** Frequencies of hydroclimate whiplash events with a 20-year recurrence interval during 2003–2023. The color scale is capped at the 99.5th percentile of the distribution of observed values to enhance visualization of spatial heterogeneity.

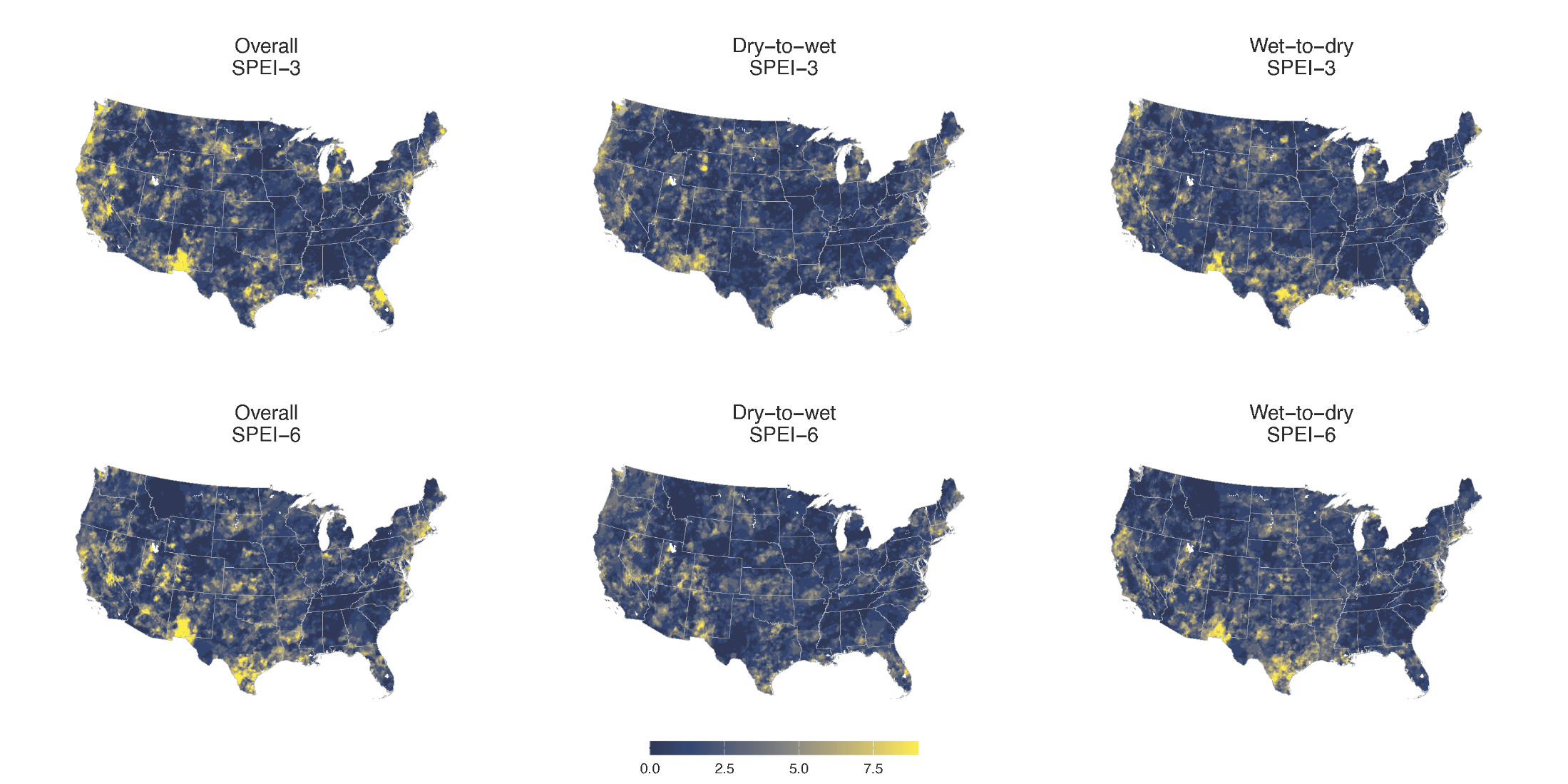

**Figure S3** Lag-response association (A) and cumulative exposure-response association over lags of 0–5 months (B) between hydroclimate whiplash events and risk of all-cause mortality by recurrence interval

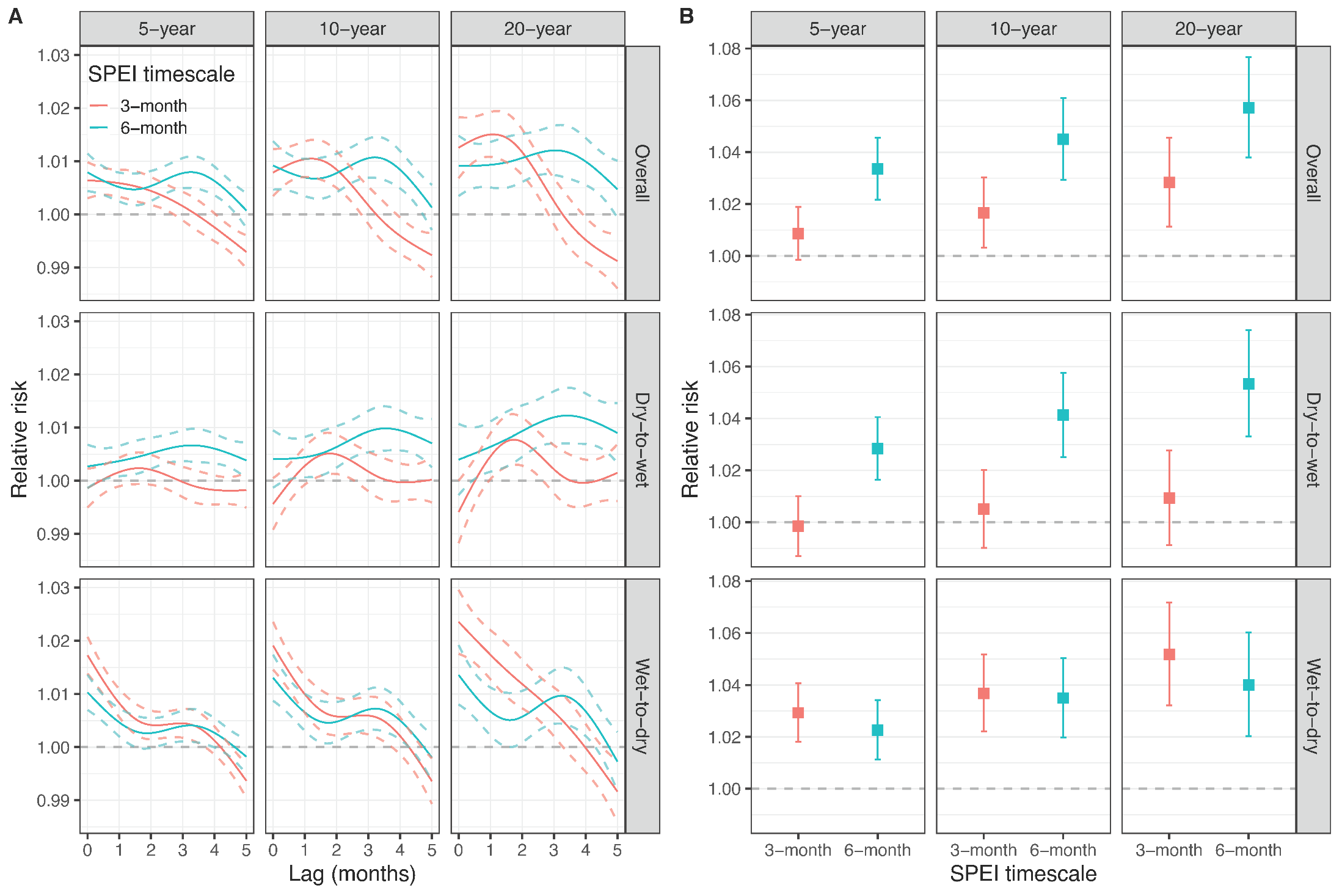

**Figure S4** Number of deaths per 100,000 population attributable to hydroclimate whiplash events across all counties in the contiguous United States during 2003–2023

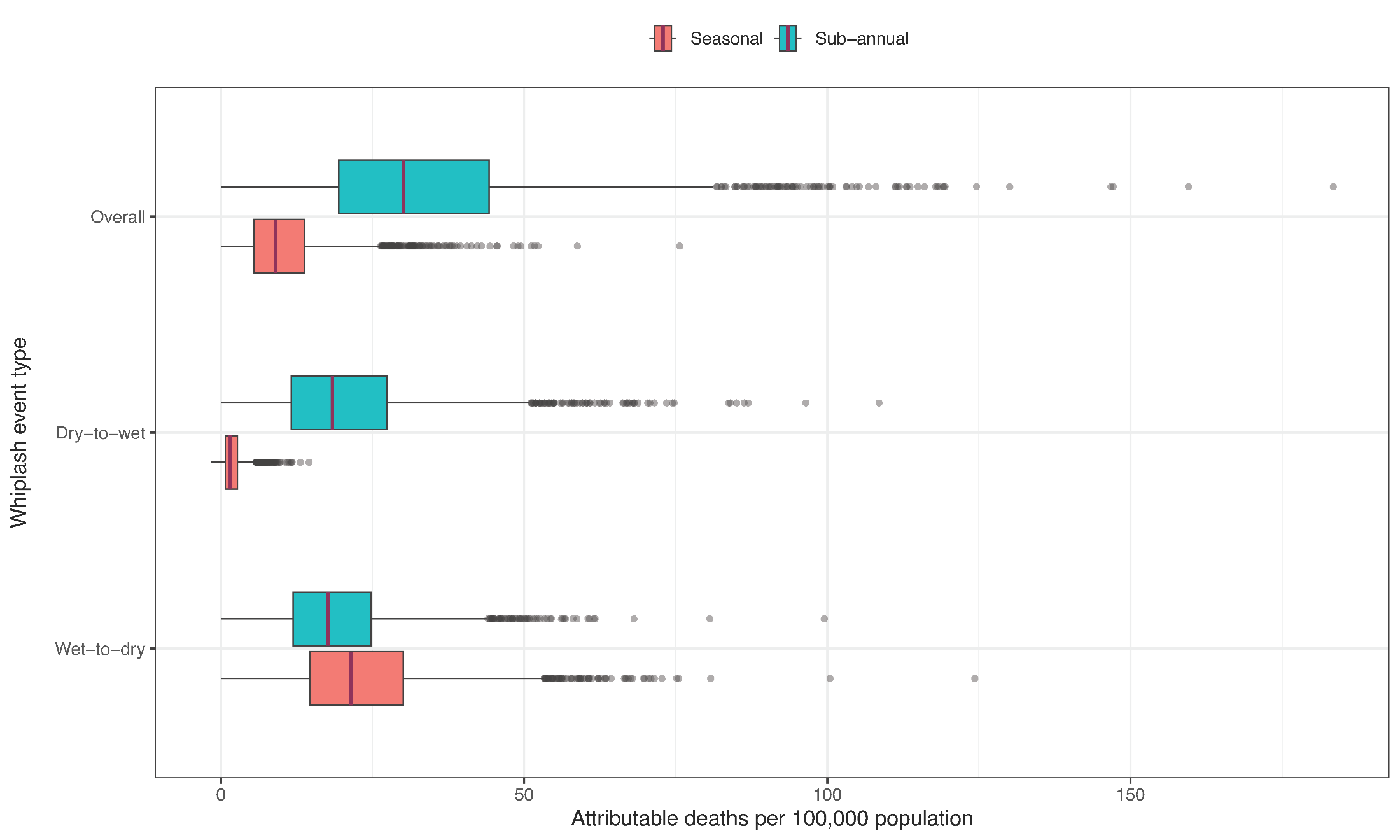

**Figure S5** Changes in annual deaths attributable to seasonal (A) and sub-annual (B) hydroclimate whiplash events from 2003–2012 to 2013–2023 per 100,000 population. Counties with red and blue borders showed a significant positive and negative trend from 2003 to 2023, respectively. The color scale is bounded between the 0.5th and 99.5th percentiles of the distribution of changes in annual attributable deaths from 2003–2012 to 2013–2023 to enhance visualization of spatial heterogeneity.

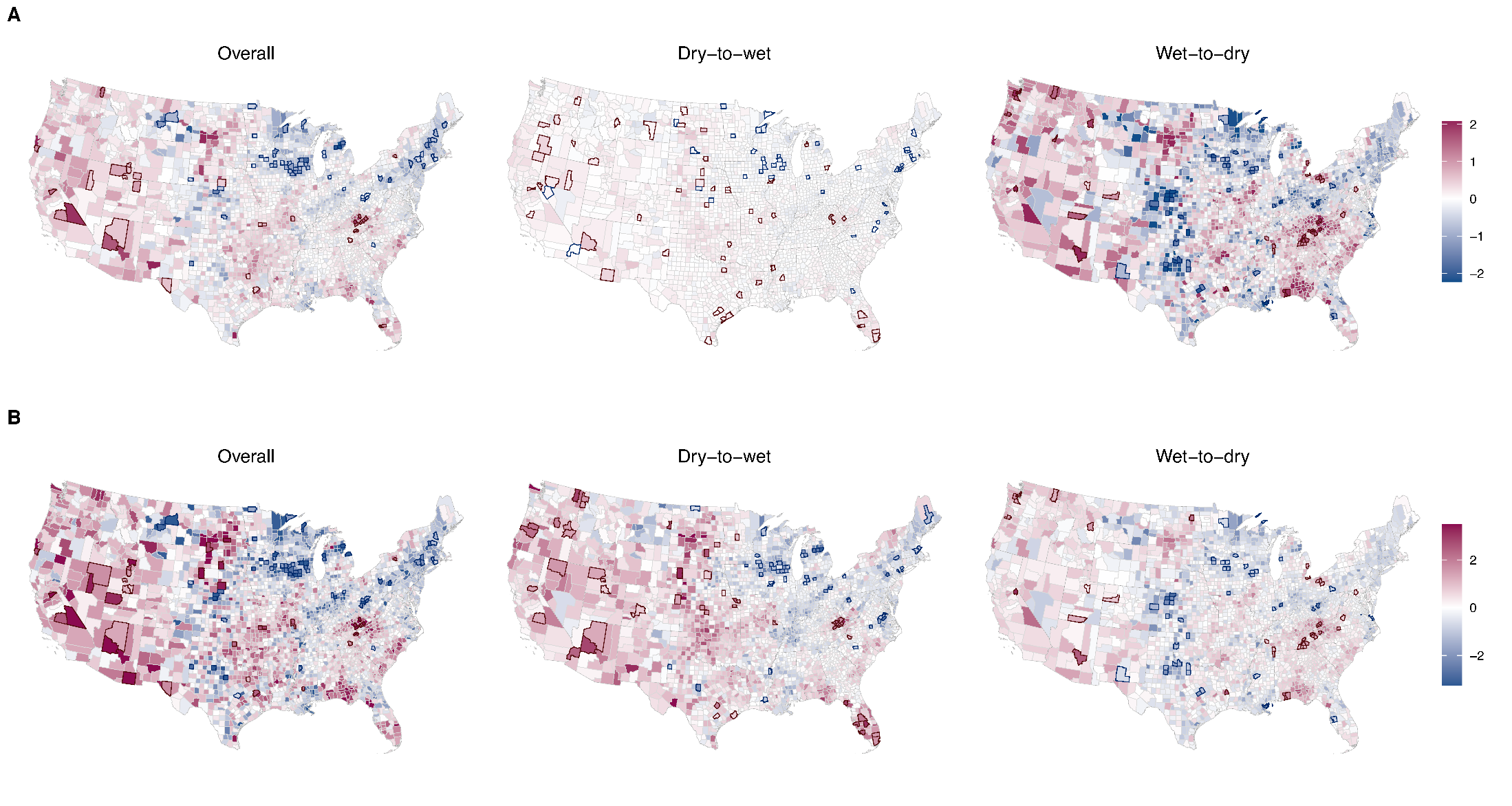

**Figure S6** Total number of deaths attributable to seasonal hydroclimate whiplash events in the contiguous United States from 2003 to 2023. Shades represent the 95% confidence intervals, derived from 5000 Monte Carlo simulations of the estimated parameters from the main model.

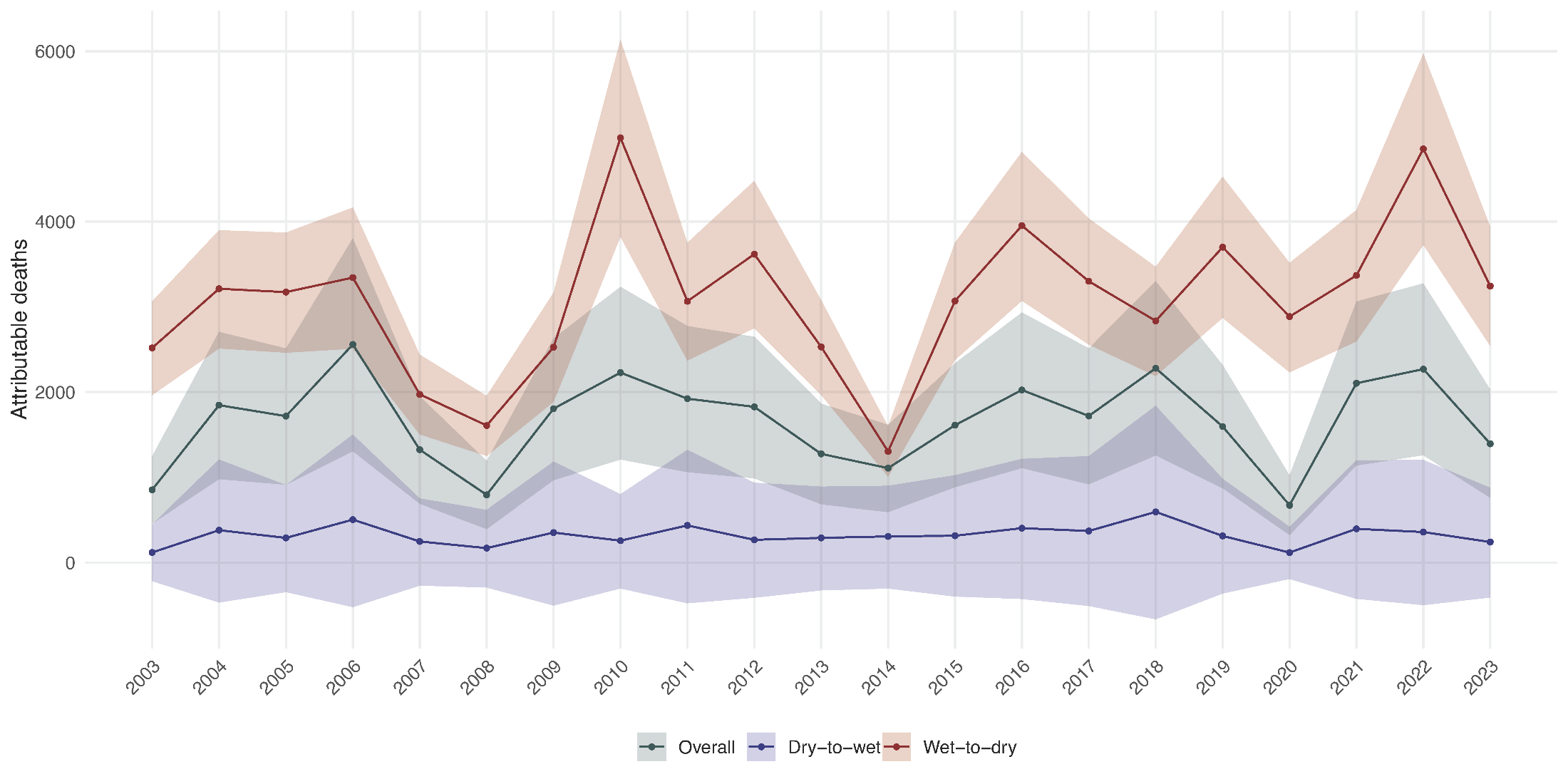

**Figure S7** Total number of deaths attributable to sub-annual hydroclimate whiplash events in the contiguous United States from 2003 to 2023. Shades represent the 95% confidence intervals, derived from 5000 Monte Carlo simulations of the estimated parameters from the main model.

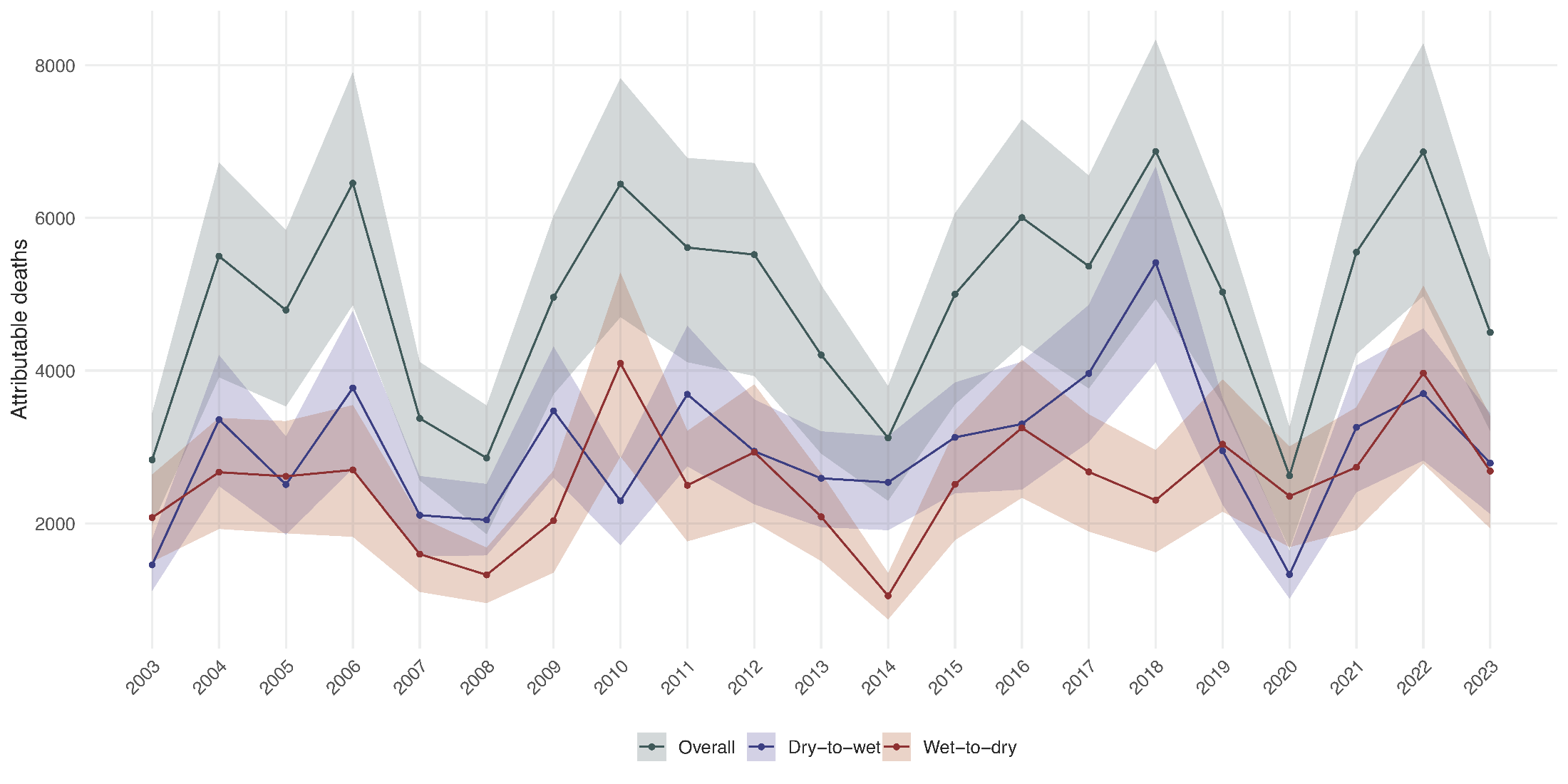

**Figure S8** Five-year moving average annual number of deaths per 100,000 population attributable to dry-to-wet hydroclimate whiplash events by state during 2003–2023. Shades represent the 95% confidence intervals, derived from 5000 Monte Carlo simulations of the estimated parameters from the main model.

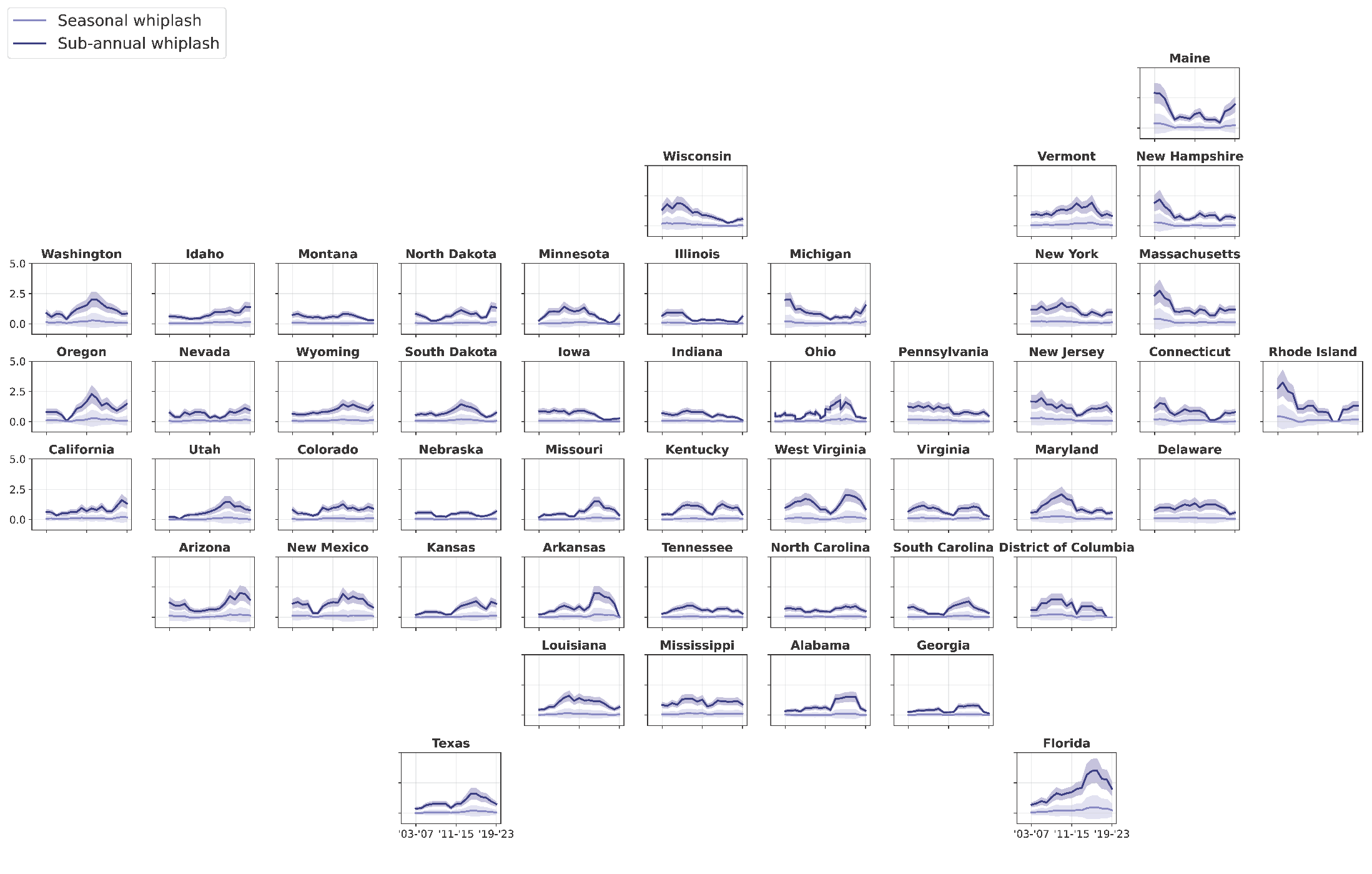

**Figure S9** Five-year moving average annual number of deaths per 100,000 population attributable to wet-to-dry hydroclimate whiplash events by state during 2003–2023. Shades represent the 95% confidence intervals, derived from 5000 Monte Carlo simulations of the estimated parameters from the main model.

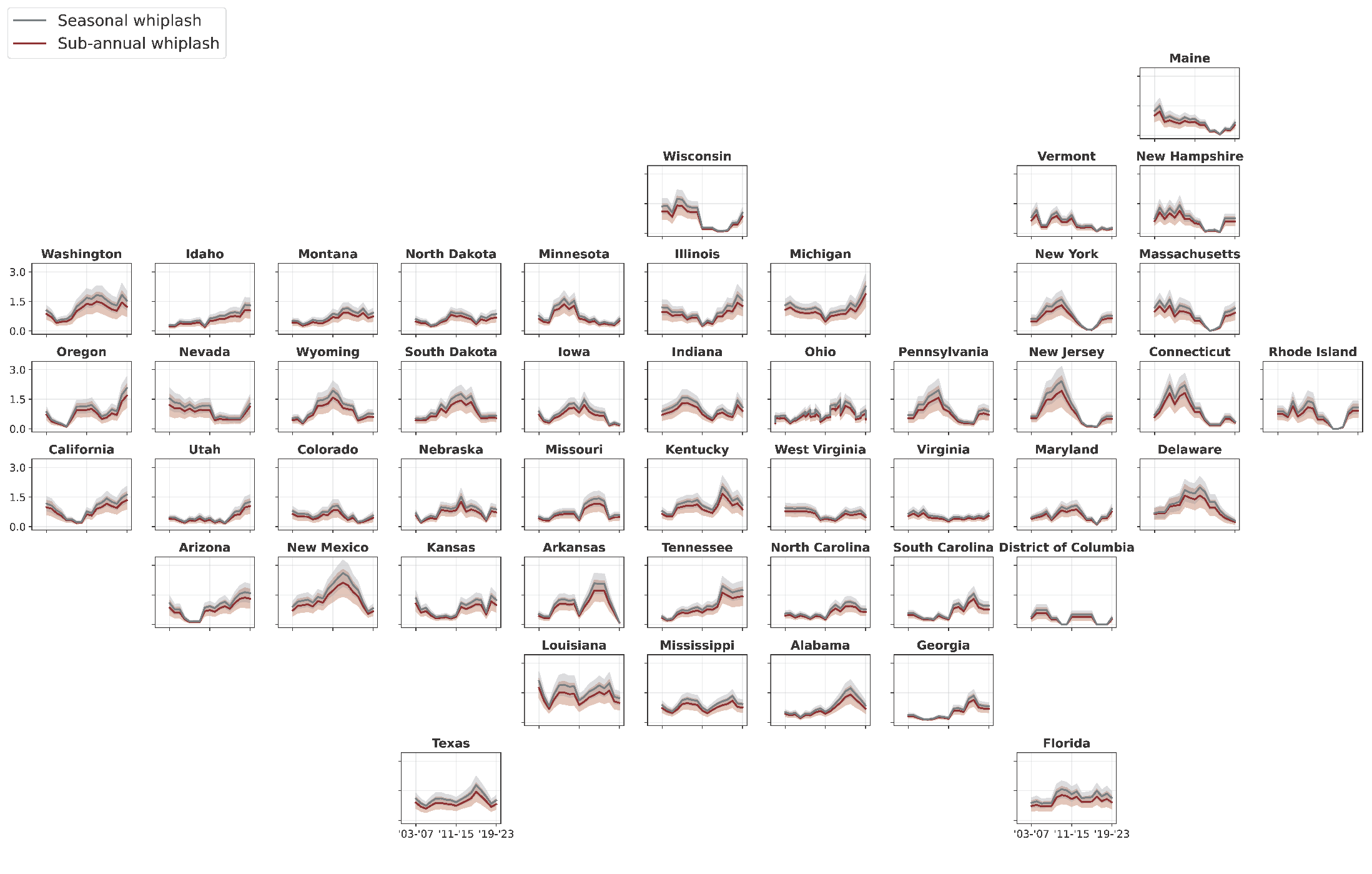

**Figure S10** Cumulative association over 0–3 months with sub-annual hydroclimate whiplash events with a 5-year recurrence interval across demographic, socioeconomic, and climate factors

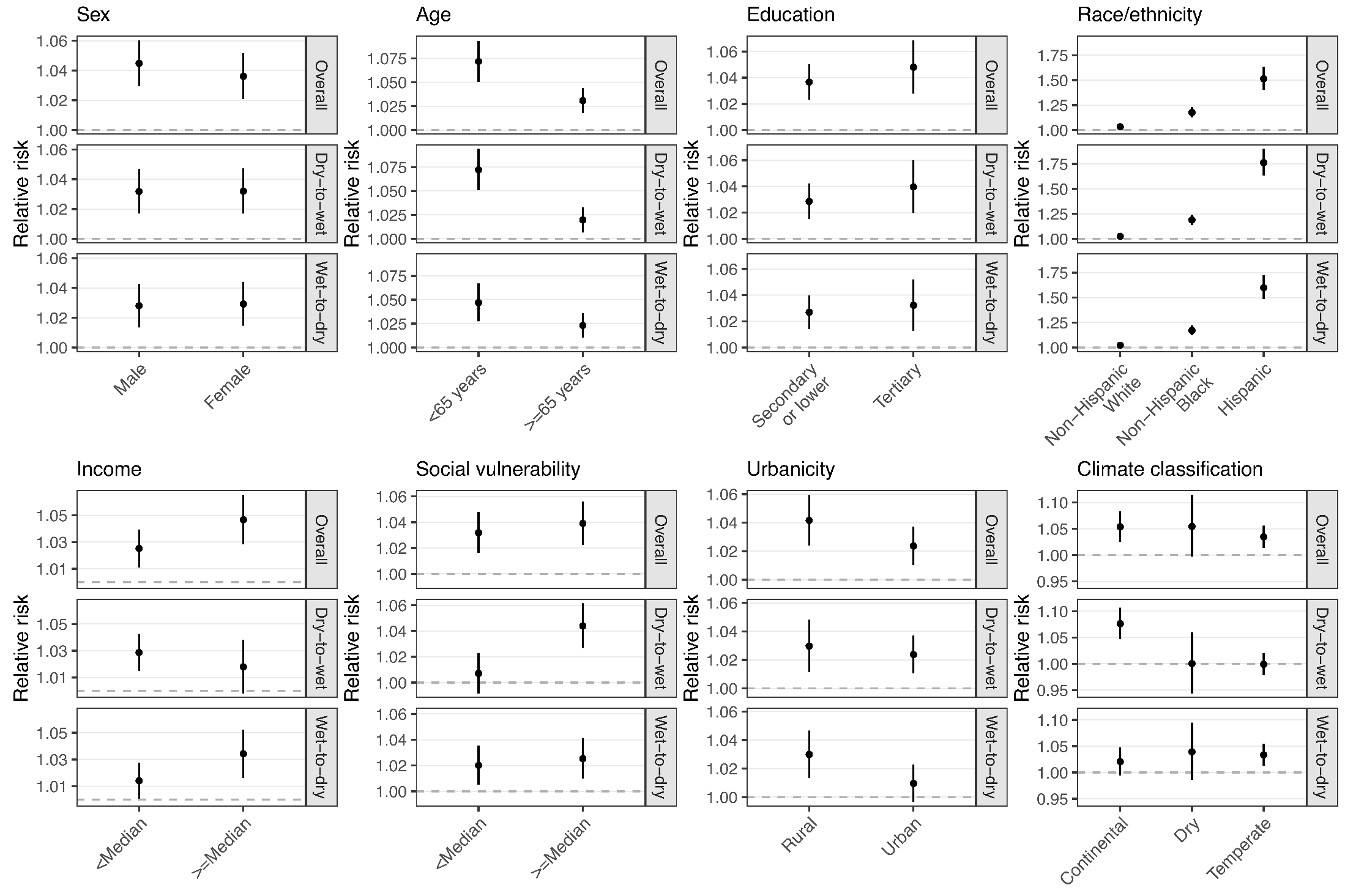
